# Parallel transmit improves 7T MRI adult epilepsy pre-surgical evaluation

**DOI:** 10.1101/2024.10.28.24316232

**Authors:** Krzysztof Klodowski, Minghao Zhang, Jian P. Jen, Daniel J Scoffings, Robert Morris, Victoria Lupson, Franck Mauconduit, Aurélien Massire, Vincent Gras, Nicolas Boulant, Christopher T. Rodgers, Thomas E. Cope

## Abstract

**Objective:** To implement parallel transmit (pTx) 7T MRI in the pre-surgical evaluation of patients with drug resistant focal epilepsy, and to compare quality and diagnostic yield to conventional single transmit (specifically, circularly polarised, CP) 7T MRI.

**Methods:** We implemented a comparative protocol comprising both pTx and CP 7T MRI in consecutive adult candidates for epilepsy surgery who had negative or equivocal 3T MRI imaging. Here we report the outcomes from the first 31 patients.

We acquired pTx and CP T_1_, T_2_, FLAIR and EDGE images, all in the same 3D 0.8mm isotropic space. 2D high-resolution T_2_ and T_2_*-weighted sequences were acquired only in CP mode due to current technological limitations.

Two neuroradiologists, a neurologist and a neurosurgeon made independent, blinded quality and preference ratings of pTx vs CP images. Quantitative methods were used to assess signal dropout.

**Results:** Blinded comparison confirmed significantly better overall quality of pTx FLAIR images (F(2,184)=13.7, p=2.88×10^-6^), while pTx MP2RAGE images were subjectively non-inferior and had improved temporal lobe coverage with quantitatively less signal drop-out.

7T-pTx revealed previously-unseen structural lesions in 9 patients (29%), confirmed 3T-equivocal lesions in 4 patients (13%), and disproved 3T-equivocal lesions in 4 patients (13%).

Lesions were better visualised on pTx than CP in 57% of cases, and never better visualised on CP.

Clinical management was altered by pTx-7T in 18 cases (58%). 9 cases were offered surgical resection and 1 LITT. 3 cases were removed from the surgical pathway because of bilateral or extensive lesions. 5 cases were offered sEEG with better targeting (in 3 because the 7T lesion was deemed equivocal by the MDT, and in 2 because the lesion was extensive).

**Significance:** Parallel transmit 7T MRI is implementable in a clinical pathway, is superior to single transmit 7T MRI, and changed management in 58% of patients scanned.

**Key points:**

1. We scanned 31 patients with parallel transmit and conventional 7T MRI, finding previously-unreported structural lesions in 9 patients (29% of cases).
2. In 13% of cases pTx 7T MRI showed that an equivocal lesion at 3T MRI was likely significant.
3. In 13% of cases pTx 7T MRI showed that an equivocal lesion at 3T MRI could be disregarded.
4. Both qualitative and quantitative quality assessments indicate superiority of pTx images over CP.
5. Future clinical implementations of 7T MRI for epilepsy should utilise parallel transmit where possible.

## Introduction

Magnetic Resonance Imaging (MRI) plays a crucial role in detecting structural lesions for presurgical planning in patients with drug-resistant epilepsy ^1^. The odds of seizure freedom after epilepsy surgery are roughly doubled if a lesion can be visualised with MRI ^2^. The International League Against Epilepsy (ILAE) Task Force’s 2021 consensus paper recommends newer Ultra-High Field (UHF) 7T MRI scanners for clinical use in this group ^3^. UHF 7T MRI offers superior spatial resolution and sensitivity compared to traditional clinical 3T MRI ^4, 5^. Importantly, 7T MRI can detect structural epileptogenic lesions that are not detectable by 3T MRI ^6^. Recent clinical results show not only superiority over 3T MRI ^7, 8^, but also correspond well with histological findings ^9^.

However, the physics of 7T MRI means that images are susceptible to dark patches – signal drop-outs in areas where the transmit B_1_ ^+^ field is weak. These dropouts commonly occur in the temporal lobes, which are a regions of particular interest for epileptogenic lesion detection ^10-12^, as well as the cerebellum. Parallel transmit (pTx) MRI can substantially mitigate these dropouts, improving the uniformity of 7T MRI images to achieve diagnostic quality throughout the whole brain ^13^. A previous limitation of pTx was the need for time-consuming per-patient calibration scans, but recent developments in plug and play, template-based Universal Pulses (UPs) allow much of the benefit of pTx at clinical timescales ^14^, at zero time penalty for the user and with no special expertise required.

In this study, we compare the performance of pTx-7T MRI against conventional, circularly polarised (CP), 7T MRI using a protocol based on the recent ILAE Task Force consensus recommendations ^3^ for the clinical assessment of patients with drug-resistant focal epilepsy. We implemented this in a ‘real-world’ epilepsy surgery pathway, scanning only those patients in whom conventional 3T MRI was inconclusive, and in whom further progress towards surgical resection would otherwise have either been impossible or required invasive stereotactic EEG (sEEG). This allows a simultaneous comparison of pTx-7T vs CP-7T and an assessment of the clinical impact of this technique.

## Methods

### Patients

We recruited 32 consecutive adult patients with drug-resistant focal epilepsy from the Cambridge Epilepsy Surgery Pathway. All patients had undergone inpatient videotelemetry (video-EEG) in which seizures had been captured, and had undertaken an FDG-PET scan. All except 2 had 3T MRI imaging that was deemed negative or equivocal for the presence of a lesion – the remaining 2 had 3T-visualied focal cortical dysplasias, but with unclear boundaries and potential impingement on dominant-hemisphere language regions, and the epilepsy multi-disciplinary team (MDT) requested 7T for better demarcation of extent. Two further patients were referred by the MDT but could not be recruited due to 7T contraindications (a tooth implant and an ear tattoo).

Patients were informed that imaging was being undertaken as part of a research study (National Research Ethics Service reference 23/WM/0008) and were informed that the scanner and sequences were not CE-marked for clinical use. All provided informed consent to participate on this basis. Clinical re-use of data as part of epilepsy surgery decision making was approved by Cambridge University Hospitals Governance and Legal structures and the UK national clinical negligence insurance scheme for trusts. A confirmation of non-objection was obtained from the UK Medicines and Healthcare Regulatory Authority.

One of the patients could not be scanned due to large body habitus (they were physically unable to fit within the scanner bore), so we report 31 datasets. The patients ranged from age 19 to age 60 (mean 34.8 years), with 15 being female and 16 male.

In four cases it was not possible to acquire the CP sequences for comparison, but in all cases we were able to complete sufficient sequences for clinical evaluation. Two patients were only able to tolerate part of the scan duration due to claustrophobia (1 patient) or having a seizure (1 patient). Two patients ran out of scanner booking time due to arriving late (1 patient) and excessive motion requiring sequence repetition (1 patient). One additional patient asked to interrupt the scan halfway through due to a feeling of claustrophobia, but after short break decided to continue and the full protocol was completed.

### MRI Acquisition

Data were acquired with a 7T MR system (MAGNETOM Terra, Siemens Healthcare, Erlangen, Germany) running VE12U SP01 software and equipped with an 8Tx/32Rx transmit/receive coil (Nova Medical, Wilmington, MA, USA). As shown in Table 1, we divided our imaging protocol into two parts. The first part comprised our candidate clinical package of parallel transmit 7T MRI sequences acquired with UP (PASTEUR package version 1.1): MP2RAGE (producing both EDGE contrast ^15, 16^, highlighting the grey-white matter border, and UNI contrast, a traditional T1-weighted image), FLAIR, and volumetric T2 TSE; as well as CP-mode T2* and high-resolution in-plane 2D T2 TSE – pTx solutions for these latter sequences are in active development but were not available in time for the commencement of this study. This clinical protocol took 40-45 minutes clock-time in total (the total duration of the sequences was 35.5 minutes, but manual shimming was performed and patient comfort checked between sequences).

**Table 1.** Sequence parameters. Previous 7T MRI epilepsy studies have used conventional single transmit (sTx) 7T MRI with the Nova 1Tx32Rx head coil. For patient comfort and to minimise motion effects, we ran all scans using the Nova 8Tx32Rx head coil. This can be run in circularly polarised (CP) mode, also known as “TrueForm” mode, which is designed to closely emulate the performance of a conventional single transmit 7T MRI scan. We have previously shown that the 8Tx32Rx head coil has slightly better SNR than the 1Tx32Rx head coil, so this choice will, if anything, underestimate the benefits of parallel transmit. Abbreviations: pTx, parallel transmit; CP, circularly polarised.

| Sequence | Transmit mode | 3D or 2D acquisition | Package | TR /ms | TE /ms | TI /ms | Voxel size / mm<sup>3</sup> | Nominal FA / ° | FOV read / mm | Slice thickness / mm | Time / min |
| --- | --- | --- | --- | --- | --- | --- | --- | --- | --- | --- | --- |
| **Proposed parallel transmit (pTx) 7T MRI clinical protocol** |  |  |  |  |  |  |  |  |  |  |  |
| MP2RAGE | pTx | 3D | PASTEUR<sup>14</sup> | 3850 | 2.27 | 800/2700 | 0.8 x<br>0.8 x<br>0.8 | 5/7 | 240 | 0.8 | 7:26 |
| FLAIR | pTx | 3D | PASTEUR | 9200 | 326 | 2300 | 0.8 x<br>0.8 x<br>0.8 | variable | 230 | 0.8 | 6:54 |
| T2 TSE | pTx | 3D | PASTEUR | 12000 | 384 | - | 0.8 x<br>0.8 x<br>0.8 | variable | 230 | 0.8 | 8:00 |
| **Control 7T scans in circularly polarised (CP) (or “TrueForm”) mode** |  |  |  |  |  |  |  |  |  |  |  |
| T2 TSE | CP | 2D | UK7T | 8870 | 77 | - | 0.48 x<br>0.48 x<br>1 | 60 | 244 | 1 | 6:32 |
| T2* | CP | 3D | UK7T | 31 | 20 | - | 0.8 x<br>0.8 x<br>0.8 | 15 | 224 | 0.8 | 7:17 |
| FLAIR | CP | 3D | Siemens | 9000 | 269 | 2600 | 0.8 x<br>0.8 x<br>0.8 | variable | 230 | 0.8 | 7:23 |
| MP2RAGE | CP | 3D | Siemens | 4300 | 1.99 | 840/2370 | 0.8 x<br>0.8 x<br>0.8 | 5/6 | 240 | 0.8 | 8:33 |
| **Parallel transmit research sequences to test feasibility in patients** |  |  |  |  |  |  |  |  |  |  |  |
| DTI | pTx | 2D | BOGAT | 6400 | 65 | - | 0.8 x<br>0.8 x<br>1.25 | 90 | 210 | 1.25 | 3:57 |

The second part comprised circularly polarised (CP, also called “TrueForm”) MP2RAGE and FLAIR scans for comparison, which are equivalent to conventional single transmit (sTx) 7T MRI. We acquired CP control scans rather than switching coils and rebooting the scanner for sTx acquisition to avoid confounding from between-scan motion, and to make the study tolerable for the patients. These sequences took a further 16 minutes, resulting in approximately one hour on scanner in total.

For some subjects, if comfort and time permitted, we last performed a diffusion tensor imaging (DTI) scan with pTx Bayesian pulse optimisation (BOGAT) ^17^. This was for sequence development and evaluation purposes, and the results were not provided to the MDT and are not reported here.

All 3D sequences were acquired in a matched 0.8mm isotropic spatial resolution and orientation to facilitate cross-sequence comparison by the neuroradiology team. Full sequence protocol PDFs to support replication at other sites are provided in Supplementary Information.

### Image evaluation - qualitative

An MRI physicist (KK) created blinded comparison images consisting of two sets of three matched orthogonal planes localised in the centre of the brain, with a slight offset for sagittal slices to ensure that brain and not falx was included. Any patient in whom there was movement artefact in either the pTx or CP sequence (more common in the latter, likely because these images were acquired later in the scan protocol when patients may have become less comfortable) was excluded from comparison. Comparison sets were randomly selected for each of the EDGE, T1-UNI, and FLAIR sequences with pTx and CP mode randomly assigned to be set ‘A’ or set ‘B’, resulting in 42 paired judgments per rater. An example is provided in Supplementary Figure 1.

Image quality was blind assessed by those members of the Cambridge multi-disciplinary team who would have clinical responsibility for their evaluation – 2 experienced neuroradiologists (JPJ and DJS), the lead neurologist (TEC) and lead neurosurgeon (RM). Raters were asked to score the images using the following 1-5 Likert scale: excellent diagnostic quality (5), good diagnostic quality (4), fair diagnostic quality (3), poor diagnostic quality (2), and non-diagnostic quality (1) ^18^.

Quality ratings were statistically compared with a single repeated measures ANOVA, implemented in Matlab 2020b, assessed for main effects of acquisition mode (pTx vs CP) and rater, as well as the within-subject factor of sequence type (FLAIR vs EDGE vs UNI), plus the interaction of rater and acquisition mode. Post-hoc paired t-tests were used to explore significant ANOVA findings.

### Image evaluation - quantitative

For a quantitative comparison of image uniformity we used Normalized Absolute Average Deviation Uniformity (NAAD) ^19^:

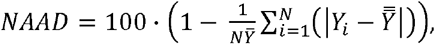

where: *Y*_*i*_ is the individual pixel value in the region of interest (ROI), 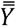 is the mean of all pixels in the ROI, and *N* is the total number of pixels in the ROI.

We calculated NAAD for white matter (WM) and grey matter (GM) separately, using the INV2 images from MP2RAGE, as these have strong WM and GM signals that display nonuniformity if present (as opposed to INV1/EDGE, which are optimised to show the grey/white boundary, and UNI/T1, which are inhomogeneity corrected).

For brain segmentation we used FreeSurfer (v6.0.0) ^20^ on UNI images from MP2RAGE acquisition. We extracted white and grey matter from INV2 images and calculated NAAD for each separately ^19^.

To determine the distribution of non-uniformity across each acquisition method, we then plotted histograms of pixel intensity, and quantified deviation from a standardised normal distribution with the Wasserstein distance ^21^ in Matlab 2020b.

### Clinical impact evaluation

Patients were referred for 7T MRI after decision by the MDT that insufficient information was present to make a surgical proposal to the patient, and the next step would otherwise be sEEG, if a plausible hypothesis could be proposed, or consideration of non-curative therapies such as vagal nerve stimulation.

After 7T imaging, patients were re-discussed by the Cambridge MDT, and where necessary also at a joint meeting of the Cambridge and King’s College Hospital MDTs (because sEEG for Cambridge patients is performed at King’s, and they also provide quantitative, MRI-coregistered PET analysis that is not available in Cambridge ^22^). As part of this meeting, the presence or absence of contributory additional information from 7T-MRI was agreed, and a consensus management decision made. A spreadsheet was kept, recording (A) what additional information was provided by 7T-MRI, if any and (B) whether the 7T-MRI changed clinical management, and if so in what way.

## Results

### Image quality assessment - qualitative

A repeated measures ANOVA with Greenhouse Geisser correction confirmed a significant main effect of Acquisition Method (pTx vs CP; F(2,184)=13.7; p=2.87×10^-6^) and a significant interaction between Rater and Acquisition Method (F(2,184)=5.92; p=0.00433). There was no significant main effect of Rater (F(2,184)=1.52; p=0.222), and a trend effect for Sequence Type (F(2,184)=2.81; p=0.0679).

Post-hoc tests demonstrated that this was because both neuroradiologists (t(11)=4.75, p=6.00×10^-4^; t(11)=4.21, p=0.0015) and the neurologist (t(11)=4.69, p=6.60×10^-4^) significantly preferred pTx FLAIR over CP FLAIR, while the neurosurgeon expressed no preference between the sequences. The neurosurgeon rated FLAIR image quality as very much higher overall (good or excellent diagnostic quality in every case for both acquisition methods), especially compared to the neuroradiologists, who provided average ratings of 2.33 and 2.42 (poor to fair diagnostic quality) for CP FLAIR and 3.75 and 3.33 (fair to good diagnostic quality) for pTx FLAIR. None of the raters displayed statistically significant rating differences for EDGE or UNI sequences (Supplementary Table 1).

### Image quality assessment - quantitative

Mean grey matter MP2RAGE INV2 NAAD of pTx images was 1.7% better (*NAAD*^*GMpTx*^=80.35%, *NAAD*_*GMCP*_ 78.62%), equivalent to an 8% reduction in non-uniformity, while white matter was 0.46% better ((*NAAD*_*WMpTx*_=86.17%, *NAAD*_*WMCP*_ 85.71%)equivalent to a 3% reduction in non-uniformity. Examination of the distribution of this non-uniformity (Figure 1) demonstrated that pixel intensity for both white matter and grey matter was much closer to a normal distribution for pTx (Wasserstein distances: WM = 0.73, GM = 0.74) than for CP (WM = 0.93, GM = 1.00). Specifically, CP images displayed a secondary peak of low pixel intensities, indicating signal drop-out, which was near-absent in the pTx images.

**Figure 1.**
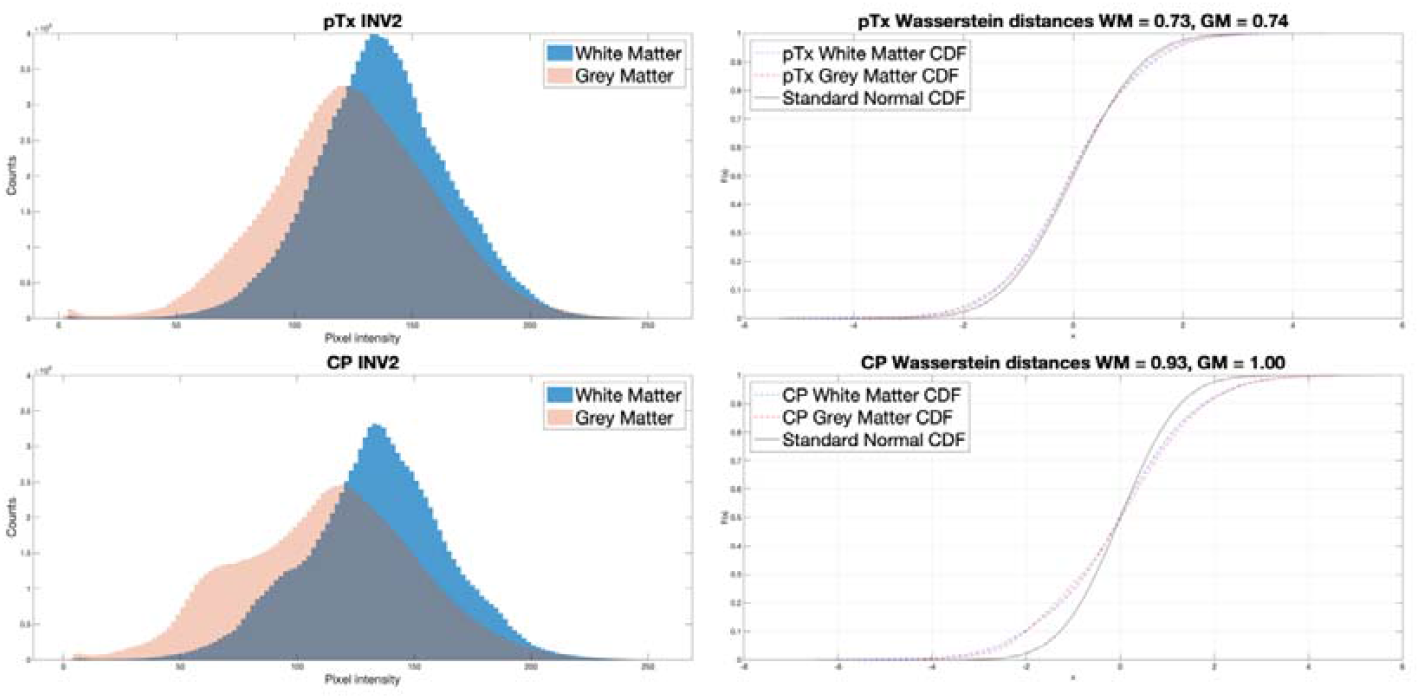
Left: Histograms of pixel intensity for the pTx and CP INV2 sequences. Right: Deviation of these distributions from a standard normal distribution, quantified by the Wasserstein distance – much higher for the CP than pTx sequences. CDF = cumulative distribution function.

### Lesion detection

7T-pTx revealed previously-unseen structural lesions in 9 patients (29%, Figure 2), confirmed 3T- equivocal lesions in 4 patients (13%), and disproved 3T-equivocal lesions in 4 patients (13%). Four patients had putative lesions detected requiring further investigation, because the MDT were not sufficiently confident they represented the epileptogenic zone (Figure 3).

**Figure 2.**
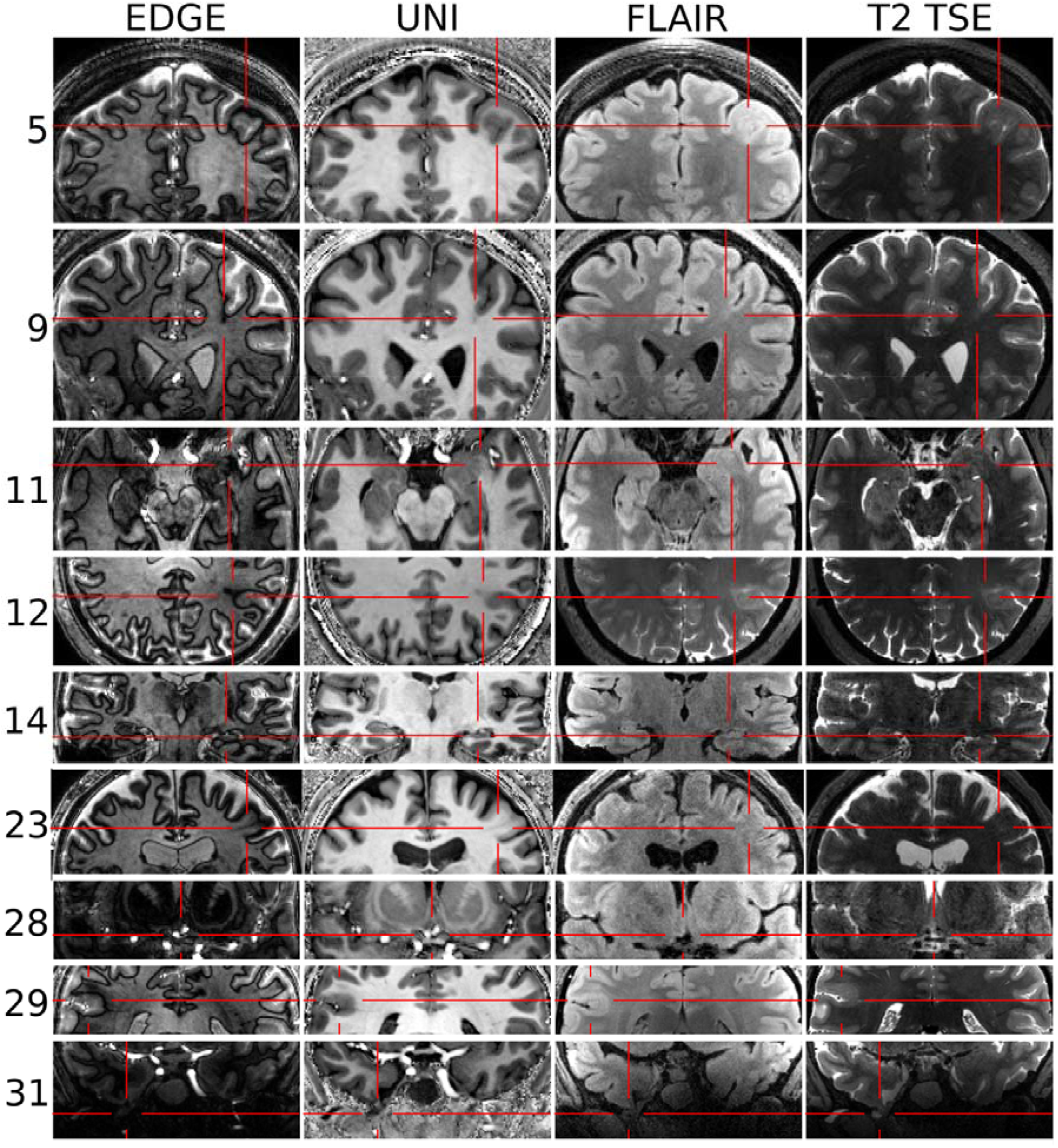
New lesions detected in first 31 patients, deemed concordant and of high confidence by the MDT. Columns show the sequences. The identified lesions in: patient 5 (FCD), patient 9 (FCD), patient 11 (amygdala enlargement), patient 12 (FCD), patient 14 (hippocampal sclerosis), patient 23 (FCD), patient 28 (dysplasia or low grade glial/glioneural lesion), patient 29 (FCD), patient 31 (right temporal encephalocele).

**Figure 3.**
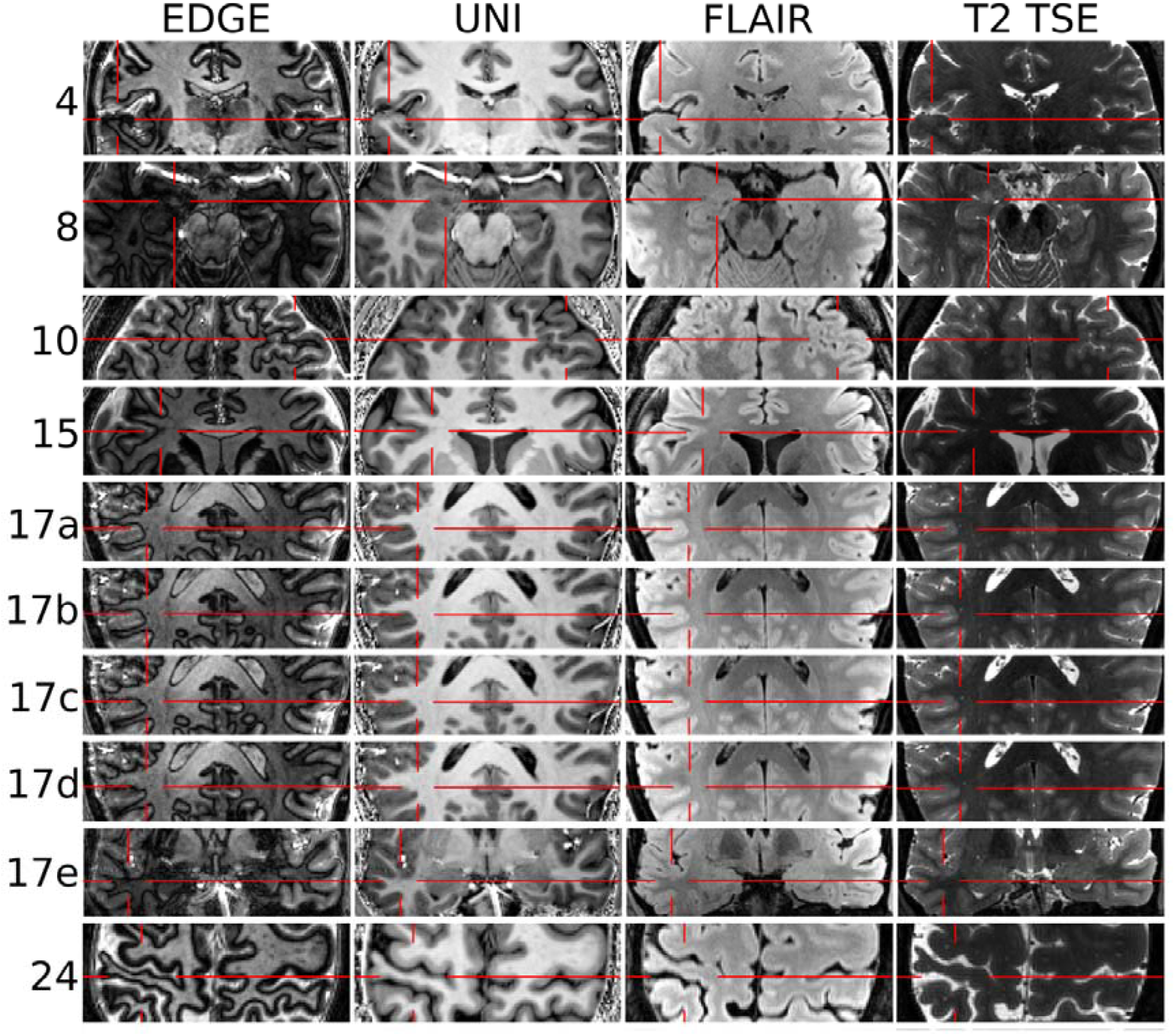
Putative lesions requiring further investigation before a surgical plan could be made. The putative lesions in: patient 4 cortical-subcortical blurring of uncertain significance, sEEG planned to evaluate significance; patient 8 likely amygdala lesion, sEEG planned to evaluate significance; patient 10 extensive polymicrogyria, sEEG planned to narrow down the epileptogenic zone; patient 15 likely FCD, ictal SPECT requested for confirmation; patient 17 rows a-d show 4 consecutive slices revealing that an equivocal lesion on 3T MRI is in fact a vessel, while row e shows a new possible FCD awaiting sEEG evaluation; patient 24: grey matter thinning likely representing perinatal infarction, sEEG planned to narrow down the epileptogenic zone.

We assessed whether new lesions were better visualised in pTx than CP mode. This was the case in 6 patients, but in one case this was due to motion artefact in the CP acquisition, and in another the CP MP2RAGE acquisition could not be completed due to patient claustrophobia. We therefore conservatively report that 4/7 (57%) of high-confidence lesions were better visualised with pTx than CP acquisition (Figures 4 and 5), there was no difference in 3/7 (43_%_), and lesions were never better visualised with CP.

**Figure 4.**
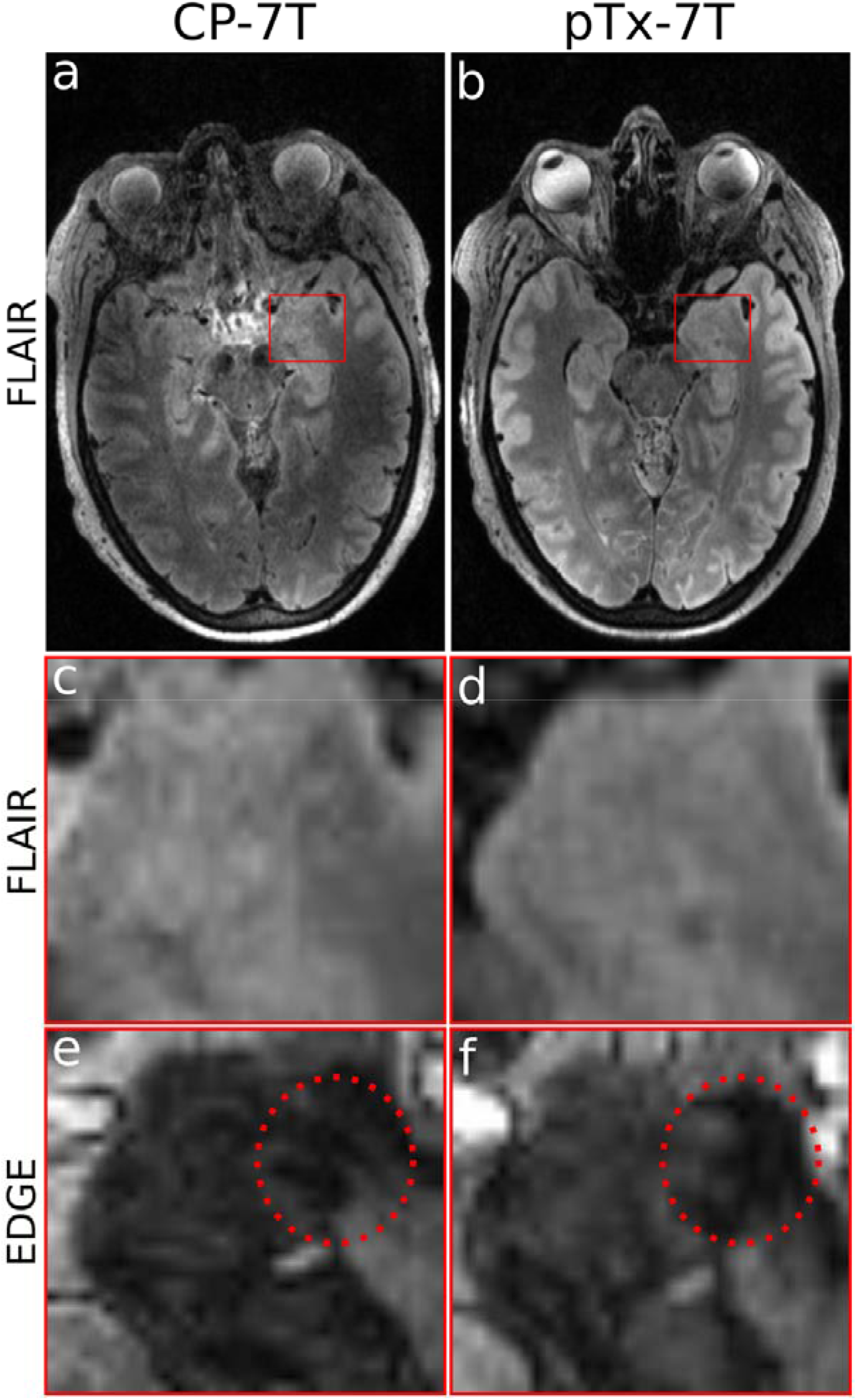
Patient 11 amygdala enlargement, visible on pTx but not CP images, due to improved uniformity of FLAIR sequence giving better definition of margins, and better clarity of EDGE contrast showing distension of lateral border.

**Figure 5.**
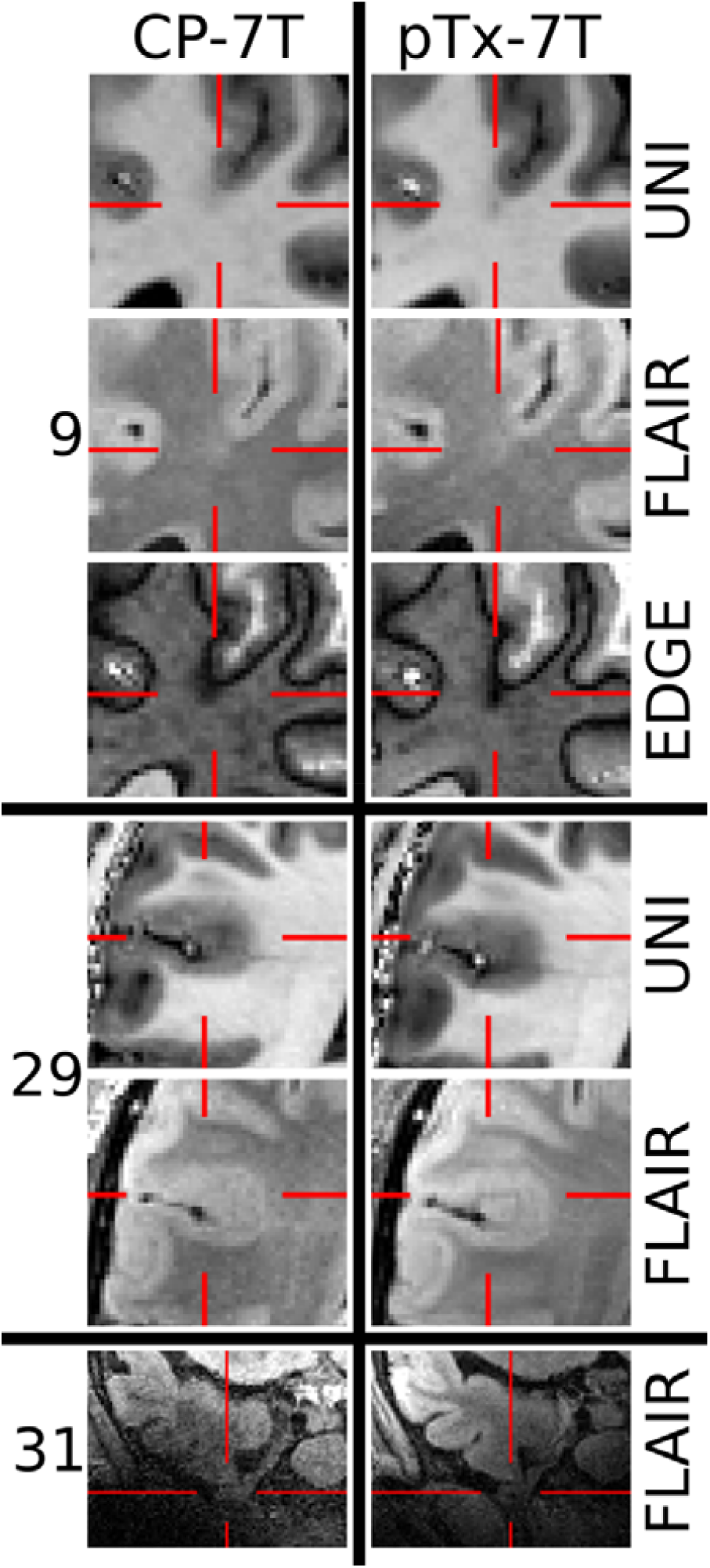
Further lesions better visualised with pTx than CP acquisitions. Patient 9: focal cortical dysplasia much more prominently displayed and crisply defined, especially in the EDGE contrast. Patient 29: focal cortical dysplasia not immediately evident on CP images, but clearly demonstrated with pTx. Patient 31: inferior temporal encephalocele falling in an area of high signal dropout and low signal to noise ratio in CP FLAIR.

### Clinical value

In 18 out of 31 patients (58%) the epilepsy MDT changed their clinical decision having evaluated the 7T MRI images. We illustrate the range and scope of this with clinical vignettes below. Overall, surgery was offered to 9 patients in whom it would not otherwise have been possible without further investigation, and Laser Interstitial Thermal Therapy (LITT) to 1 patient. 3 cases were removed from the surgical pathway because of bilateral or extensive lesions. 5 cases were offered sEEG with better targeting (in 3 because the 7T lesion was deemed equivocal by the MDT, and in 2 because the lesion was extensive). 1 additional patient received a different operation to that originally proposed in part because 7T excluded a 3T-equivocal lesion (this was confirmed by negative sEEG findings in the 3T- implicated area, and after left temporal lobectomy the patient was found to have hippocampal sclerosis that was not visible at either 3T or 7T).

### Case Vignettes

#### Patients where surgery offered (Figure 2)

Patient 5, female, age 31-35. 7T demonstrates a left superior frontal cortex dysplasia, not visible at 3T, which was concordant with PET hypometabolism. The patient was offered a choice between surgery with awake craniotomy and electrocorticography for language or sEEG, but this has been deferred due to improved seizure control on increased anti-seizure medications.

Patient 9, female, age 26-30. 7T demonstrates focal cortical dysplasia of left superior frontal sulcus with transmantle sign, concordant with PET. Previous 3T had demonstrated a malrotated hippocampus, but 7T demonstrated good hippocampal structure and no associated FCD. Patient offered and accepted surgery, but has deferred the date until the end of her University Masters degree.

Patient 11, female, age 51-55. 7T demonstrates new left amygdala enlargement, better visualised on pTx than CP sequences (Figure 4). Surgery likely to be offered, but awaiting quantitative PET analysis for confirmation of concordance.

Patient 12, female, age 41-45. Patient already known to have left parietal focal cortical dysplasia at 3T, but margins poorly defined and potential impingement on language areas. 7T delineated the lesion very precisely. Patient offered surgery with awake craniotomy and electrocorticography for language.

Patient 14, male, age 41-45. 7T demonstrates new left hippocampal sclerosis, concordant with EEG. Surgery offered.

Patient 23, male, age 56-60. 3T demonstrated likely focal cortical dysplasia in frontal eye fields of unclear extent, confirmed and much more precisely demonstrated at 7T. Surgery with intra-operative functional mapping offered.

Patient 28, male, age 21-25. 7T demonstrates small lesion in right paraterminal gyrus, likely representing dysplasia or a low grade glial/glioneuronal lesion. LITT offered due to lesion size and deep location near hypothalamus.

Patient 29, male, age 16-20. 3T equivocal right supramarginal gyrus dysplasia confirmed and well demarcated at 7T. Surgery with intra-operative functional mapping offered.

Patient 31, female, age 21-25. 7T demonstrates right temporal meningocele, concordant with PET hypometabolism. Surgery likely to be offered, pending neuropsychology assessment.

#### Patients referred for targeted sEEG or other investigation (Figure 3, all results awaited)

Patient 4, female, age 36-40: 7T initially reported as normal, but at MDT review the right anterior mid-temporal EEG onset was thought to have a potential correlate in cortical-subcortical blurring in right superior temporal gyrus (STG) – patient listed for sEEG with targeting of the putative lesion

Patient 8, female, age 36-40. 7T demonstrates a likely right amygdala lesion. Patient offered targeted sEEG because of bilateral EEG changes, negative PET, and non-lateralising semiology.

Patient 10, male, age 16-20. 7T demonstrated polymicrogyria in the left anterior frontal lobe with associated PET abnormality. Because the lesion is quite extensive, and close to language regions, sEEG has been offered.

Patient 15, female, age 46-50. 3T demonstrated two equivocal lesions in right frontal lobe. 7T recapitulated one lesion, and excluded the other lesion. PET was normal, and there was much debate at MDT about whether sufficient information was present to offer surgery directly. Ultimately, out of caution, it was decided to perform ictal SPECT (result awaited) – if this is concordant, surgery will be offered, otherwise sEEG will be performed with better targeting.

Patient 17, female, age 16-20. 3T equivocal lesion in right parietal lobe demonstrated to be a vessel with thin cut 7T EDGE sequence. 7T demonstrates new equivocal lesion with PET concordance in right superior temporal gyrus, but videotelemetry discordant, so patient listed for targeted sEEG.

Patient 24, female, age 31-35. 7T demonstrates cortical thinning in the right post-central gyrus, likely representing a peri-natal infarction. Concordant with EEG and thought causative, but sEEG offered because of large lesion size and the potential for functional deficit.

### Incorrect surgery avoided

Patient 1, female, age 21-25. Presumptive 3T lesion demonstrated to be venous spaces at 7T. Because of this, instead of proceeding directly to operative management the patient had sEEG. This confirmed an absence of abnormality in the 3T-reported lesion, and instead implicated the left medial temporal lobe. A left temporal lobectomy was performed and histology confirmed hippocampal sclerosis. Even in retrospect, we do not think that this is visually evident at 3T or 7T, although quantitative analyses are in development.

Patient 6, male, age 46-50. 3T equivocal lesion in right insula, which was concordant with video-EEG, demonstrated by 7T to be a peri-vascular space. Patient removed from surgical pathway due to lack of a clear hypothesis for sEEG.

Patient 16, female, age 21-25. Patient known to have right polymicrogyria at 3T, but 7T requested because of some suspicion of additional left hippocampal sclerosis. In fact, 7T demonstrated additional left sided polymicrogyria, so the patient was deemed ineligible for surgery due to significant bilateral lesions, and vagal nerve stimulator was offered instead.

## Discussion

This study tested the feasibility and impact of implementing a parallel transmit (pTx) version of the ILAE Consensus recommended 7T MRI protocols for epilepsy ^3^ in a real-world clinical epilepsy surgery pathway. We directly evaluated the quality of pTx vs CP sequences and found pTx to be both qualitatively and quantitatively superior. We demonstrated that pTx-7T influenced clinical management in 58% of cases, despite our patients having been selected because of negative or equivocal state-of-the-art imaging at 3T.

We demonstrated that pTx-7T was feasible to implement, with less than 10% of patients being excluded due to 7T contraindications, and no cases of patients unable to complete a clinically diagnostic set of sequences due to discomfort. Patient engagement activities undertaken after the study reported a high level of tolerability from patients, and only minor and occasional negative experiences such as dizziness on scanner entry and additional claustrophobia from the head coil. One patient experienced a seizure during their scan, but this was well managed by our radiography staff, who removed the patient from the scanner and made them safe, with high patient satisfaction afterwards, especially when we told them that sufficient data had been obtained for clinical evaluation.

Detection of new lesions in 29% of scanned patients, and clarification of 3T equivocal lesions in a further 26% (with a 50/50 split between verified and excluded) is more encouraging than a recent study by Hangel et al., who implemented the task force consensus protocols in single transmit mode, finding new lesions in 19% of cases ^12^. This increased yield perhaps reflects our finding that 57% of lesions were better demonstrated in pTx than CP acquisitions.

In the majority of case series of epileptogenic lesions, the most common abnormality is hippocampal sclerosis, however in our study this represented only 1 of 13 detected lesions. There are two likely reasons for this. The first is that the incidence of hippocampal sclerosis has been observed to be decreasing over decades ^23^, likely as a result of lower incidences of childhood infection. The second is that hippocampal sclerosis is usually well detected at lower field strengths, and all of the patients in this study had negative or equivocal 3T MRI as an inclusion criterion. Although, with this in mind, it is interesting to note that our cohort included one patient who had demonstrated hippocampal sclerosis in post-operative histopathology that was not demonstrated, even in retrospect, at 3T or 7T.

Conversely, our study demonstrated a high proportion of malformations of cortical development, especially focal cortical dysplasias. These lesions are often small and subtle, being invisible or equivocal at lower field strengths. The high resolution of 7T acquisition, especially in combination with the EDGE contrast that emphasises the grey/white matter border ^15, 16^, brings significant additional power to the detection of these epileptogenic lesions. Our subjective experience was that quantitative, co-registered FDG-PET was particularly helpful in guiding the neuroradiological assessment, demonstrating concordance, and increasing lesion-reporting confidence.

### Limitations

Because of the delays inherent in performing neurosurgical resections and sEEG, the primary limitation of our study is that it reports only lesion visualisation and MDT outcomes. More robust and objective outcomes are not yet available, but will be reported in subsequent papers reporting sEEG- 7T concordance, histopathological data from resected 7T lesions, and ultimately patient seizure freedom and quality of life. Our study was also only at a single centre – we are currently developing a multi-site extension.

There was undoubtedly a learning curve over the course of this study, with neuroradiologists becoming more familiar with 7T images, and clinicians in the MDT becoming more confident in relying on them with increasing experience. Naturally there was some initial scepticism about relying on a technique that does not yet have CE-marking or multi-centre evaluation, and this may have led to some conservative decisions to proceed to additional investigation, especially for patients scanned early in the series. We also cannot exclude the possibility of bias, with clinicians involved in the study also being part of the MDT making decisions, although they always represented a minority of attendees and were very aware of their responsibilities to patient safety.

All of our clinical assessments relied on manual scan reads, informed at the MDT meeting by information from seizure semiology, video-EEG, PET-determined metabolism, and neuropsychological evaluation. There is evidence that quantitative and automated image analysis methods can improve the sensitivity and confidence of lesion detection ^24, 25^. These methods have not yet been validated in, or adapted to, 7T images, and this will be a focus of our future work.

Our comparison images were acquired in CP mode, which uses the pTx coil to mimic sTx acquisition, but does not require coil swapping. We have shown previously that CP acquisition using Nova 8Tx head coil is equivalent to Nova 1Tx head coil acquisition, apart from the better SNR achieved with the former ^26, 27^. This means that any superiority of the pTx acquisition over CP is at least the same or better as the comparison with sTx acquisition using the standard Nova 1Tx head coil.

It is important to recognise that, although our pTx images were superior to CP in qualitative and quantitative image quality, they were not perfect, especially in inferior temporal lobe. One patient with a 3T-demonstrated inferior temporal meningocele, being scanned to ascertain whether there was an associated cortical dysplasia, had 7T images entirely obscured by drop-out in the relevant area with both pTx and CP. Similarly, the EDGE contrast frequently defined the grey/white border less clearly in temporal lobes than elsewhere. Work is ongoing to improve these issues. Similarly, we and others are developing pTx methods for 2D high-resolution T_2_ acquisitions and susceptibility weighted imaging, to allow a full suite of pTx sequences for epilepsy evaluation. At present we would recommend 7T MRI as next-stage investigation only after 3T MRI, not as a direct replacement.

## Conclusion

We conclude that pTx-7T MRI is not inferior to sTx-MRI in any case, and produces images of qualitatively and quantitatively superior quality. We therefore recommend that it should be adopted as the primary method for 7T MRI for epilepsy.

We demonstrate that the introduction of pTx-7T MRI to a real-world epilepsy surgery pathway changed clinical management for more than half of the patients scanned, making it highly likely that it will be cost-effective, especially given its low cost in comparison to sEEG, as well as the health-economic burdens of intractable epilepsy.

## Data Availability

Scan data are available on reasonable request but not publicly available due to patient confidentiality.

## Abbreviations

CP: Circularly Polarized acquisition (equivalent to sTx using the parallel transmit coil)
DTI: Diffusion Tensor Imaging
EDGE: Edge-Enhancing Gradient Echo (first inversion image from MP2RAGE)
FCD: Focal Cortical Dysplasia
FLAIR: Fluid Attenuated Inversion Recovery
ILAE: International League Against Epilepsy
MDT: Multi Disciplinary Team
NAAD: Normalised Absolute Average Deviation
pTx: parallel transmit acquisition
sEEG: stereotactic Electroencephalography
sTx: single transmit acquisition
UHF: Ultra High Field
UNI: MP2RAGE uniform image

## Acknowledgements

We thank members of the Cambridge University Hospitals and King’s College Hospital Epilepsy Multidisciplinary Teams, and especially the neurophysiology leads Rachel Thornton and Nandini Mullatti, for their careful consideration of the 7T MRI findings and for taking the time to formally document these discussions.

We thank the radiographers at the Wolfson Brain Imaging Centre, who provided tireless support to the patients and researchers throughout this study.

This work was supported by the Cambridge University Hospitals Academic Fund for functional neurosurgery and the University of Cambridge’s Medical Research Council Impact Acceleration Account Award (UKRI Grant Reference - MR/X502844/1).

MZ was supported by an MRC PhD studentship (MR N013433-1).

This study was supported by the NIHR Cambridge Biomedical Research Centre (NIHR203312) and an MRC Clinical Research Infrastructure Award for 7T (MR/M008983/1). The views expressed are those of the authors and not necessarily those of the NIHR or the Department of Health and Social Care. For the purpose of open access, the authors have applied a CC BY public copyright licence to any Author Accepted Manuscript version arising from this submission.

## Supplementary material

**Supplementary figure 1:**
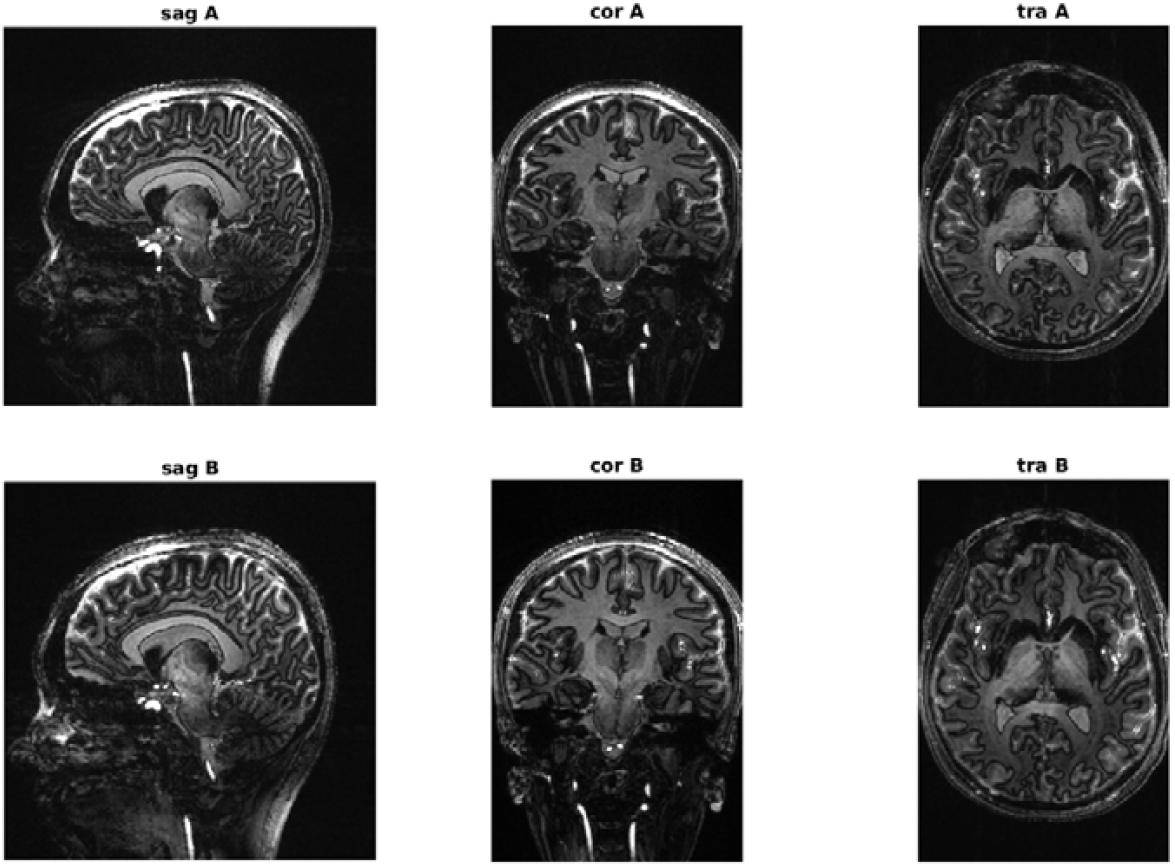
An example comparison set for qualitative pTx vs CP evaluation.

**Supplementary Table 1.**
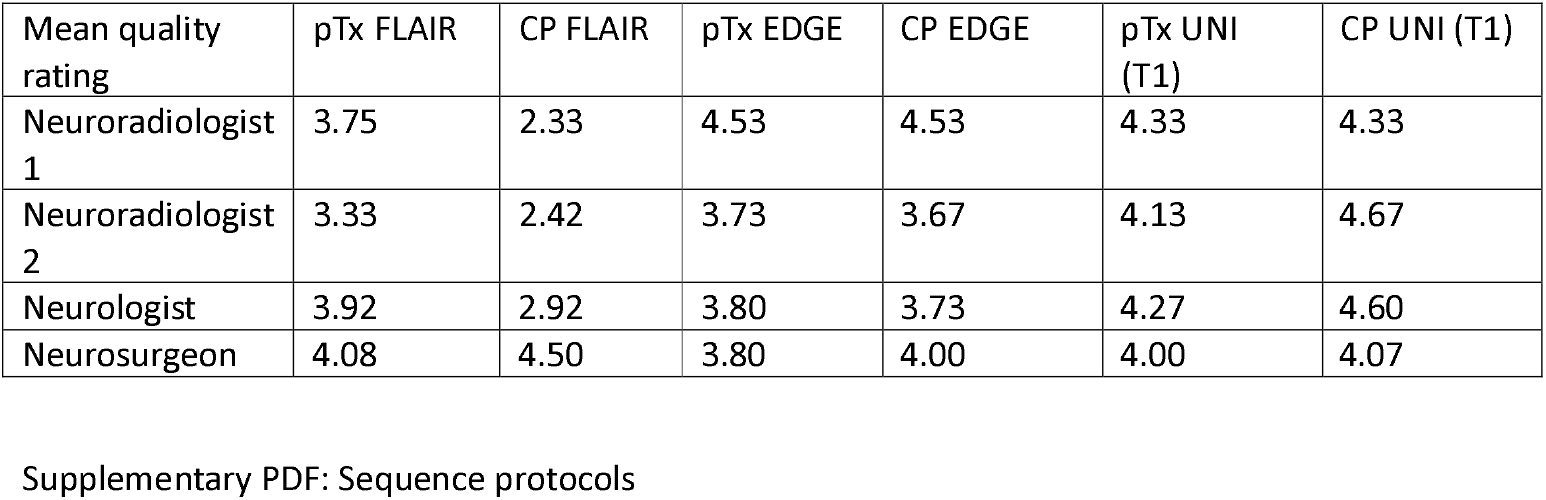
Image quality blind-assessed by two radiologists (JPJ, DJS), a neurologist (TEC) and a neurosurgeon (RM), using a standardised 1-5 Likert scale : excellent diagnostic quality (5), good diagnostic quality (4), fair diagnostic quality (3), poor diagnostic quality (2), and non-diagnostic quality (1).

| Mean quality rating | pTx FLAIR | CP FLAIR | pTx EDGE | CP EDGE | pTx UNI (T1) | CP UNI (T1) |
| --- | --- | --- | --- | --- | --- | --- |
| Neuroradiologist 1 | 3.75 | 2.33 | 4.53 | 4.53 | 4.33 | 4.33 |
| Neuroradiologist 2 | 3.33 | 2.42 | 3.73 | 3.67 | 4.13 | 4.67 |
| Neurologist | 3.92 | 2.92 | 3.80 | 3.73 | 4.27 | 4.60 |
| Neurosurgeon | 4.08 | 4.50 | 3.80 | 4.00 | 4.00 | 4.07 |

