## Supplementary Materials for "Parallel transmit improves 7T MRI adult epilepsy pre-surgical evaluation"

**\\USER\WBIC Protocols Ready\Protocol 569\P00569_Epilepsy_20230628\AAHead_Scout_32ch-head-coil**

TA: 0:18 PM: REF Voxel size: 1.6×1.6×1.6 mmPAT: 3 Rel. SNR: 1.00 : fl

### Properties

| | |
|---|---|
| Prio recon | Off |
| Load images to viewer | On |
| Inline movie | Off |
| Auto store images | On |
| Load images to stamp segments | Off |
| Load images to graphic segments | On |
| Auto open inline display | Off |
| Auto close inline display | Off |
| Start measurement without further preparation | Off |
| Wait for user to start | Off |
| Start measurements | Single measurement |

### Routine

| | |
|---|---|
| Slab group | 1 |
| Slabs | 1 |
| Dist. factor | 20 % |
| Position | L0.0 A0.5 H16.3 mm |
| Orientation | Sagittal |
| Phase enc. dir. | A >> P |
| Phase oversampling | 0 % |
| Slice oversampling | 0.0 % |
| Slices per slab | 128 |
| FoV read | 260 mm |
| FoV phase | 100.0 % |
| Slice thickness | 1.6 mm |
| TR | 4.17 ms |
| TE | 1.53 ms |
| Averages | 1 |
| Concatenations | 1 |
| Filter | B1 filter |
| Coil elements | AC |

### Contrast - Common

| | |
|---|---|
| TR | 4.17 ms |
| TE | 1.53 ms |
| Flip angle | 16 deg |

### Contrast - Dynamic

| | |
|---|---|
| Averages | 1 |
| Averaging mode | Short term |
| Reconstruction | Magnitude |
| Measurements | 1 |

### Resolution - Common

| | |
|---|---|
| FoV read | 260 mm |
| FoV phase | 100.0 % |
| Slice thickness | 1.6 mm |
| Base resolution | 160 |
| Phase resolution | 100 % |
| Slice resolution | 69 % |
| Phase partial Fourier | 6/8 |
| Slice partial Fourier | 6/8 |
| Trajectory | Cartesian |

### Resolution - iPAT

| | |
|---|---|
| PAT mode | GRAPPA |
| Accel. factor PE | 3 |
| Ref. lines PE | 24 |

### Resolution - iPAT

| | |
|---|---|
| Accel. factor 3D | 1 |
| Reference scan mode | Integrated |

### Resolution - Filter Image

| | |
|---|---|
| Image Filter | Off |
| Distortion Corr. | Off |
| Prescan Normalize | Off |
| Normalize | Off |
| B1 filter | On |
| Unfiltered images | Off |

### Resolution - Filter Rawdata

| | |
|---|---|
| Raw filter | Off |
| Elliptical filter | Off |

### Geometry - Common

| | |
|---|---|
| Slab group | 1 |
| Slabs | 1 |
| Dist. factor | 20 % |
| Position | L0.0 A0.5 H16.3 mm |
| Orientation | Sagittal |
| Phase enc. dir. | A >> P |
| Slice oversampling | 0.0 % |
| Slices per slab | 128 |
| FoV read | 260 mm |
| FoV phase | 100.0 % |
| Slice thickness | 1.6 mm |
| TR | 4.17 ms |
| Multi-slice mode | Sequential |
| Series | Ascending |
| Concatenations | 1 |

### Geometry - AutoAlign

| | |
|---|---|
| Slab group | 1 |
| Position | L0.0 A0.5 H16.3 mm |
| Orientation | Sagittal |
| Phase enc. dir. | A >> P |
| Initial Position | Isocenter |
| L | 0.0 mm |
| P | 0.0 mm |
| H | 0.0 mm |
| Initial Rotation | 0.00 deg |
| Initial Orientation | Transversal |

### Geometry - Tim Planning Suite

| | |
|---|---|
| Set-n-Go Protocol | Off |
| Table position | H |
| Table position | 0 mm |
| Inline Composing | Off |

### System - Miscellaneous

| | |
|---|---|
| Positioning mode | REF |
| Table position | H |
| Table position | 0 mm |
| MSMA | S - C - T |
| Sagittal | R >> L |
| Coronal | A >> P |
| Transversal | F >> H |
| Coil Combine Mode | Adaptive Combine |

### System - Miscellaneous

| | |
|---|---|
| Save uncombined | Off |
| Matrix Optimization | Off |
| Coil Select Mode | Default |

### System - Adjustments

| | |
|---|---|
| B0 Shim mode | Tune up |
| B1 Shim mode | TrueForm |
| Confirm freq. adjustment | Off |
| Assume Dominant Fat | Off |
| Assume Silicone | Off |
| Adjustment Tolerance | Auto |

### System - Adjust Volume

| | |
|---|---|
| Position | Isocenter |
| Orientation | Transversal |
| Rotation | 0.00 deg |
| A >> P | 263 mm |
| R >> L | 350 mm |
| F >> H | 350 mm |
| Reset | Off |

### System - pTx Volumes

| | |
|---|---|
| B1 Shim mode | TrueForm |
| Excitation | Non-sel. |

### System - Tx/Rx

| | |
|---|---|
| Frequency 1H | 297.231131 MHz |
| Correction factor | 1 |
| Gain | High |
| Img. Scale Cor. | 1.000 |
| Reset | Off |
| ? Ref. amplitude 1H | 0.000 V |

### Physio - PACE

| | |
|---|---|
| Resp. control | Off |
| Concatenations | 1 |

### Inline - Common

| | |
|---|---|
| Flip angle | 16 deg |
| Measurements | 1 |
| Time to center | 7.7 s |

### Inline - Inline

| | |
|---|---|
| Subtract | Off |
| Measurements | 1 |
| StdDev | Off |
| Save original images | On |

### Inline - MIP

| | |
|---|---|
| MIP-Sag | Off |
| MIP-Cor | Off |
| MIP-Tra | Off |
| MIP-Time | Off |
| Save original images | On |

### Inline - Composing

| | |
|---|---|
| Inline Composing | Off |
| Distortion Corr. | Off |

### Inline - MapIt

| | |
|---|---|
| Save original images | On |
| MapIt | None |
| Flip angle | 16 deg |
| Measurements | 1 |
| Contrasts | 1 |
| TR | 4.17 ms |
| TE | 1.53 ms |

### Sequence - Part 1

| | |
|---|---|
| Introduction | On |
| Dimension | 3D |
| Asymmetric echo | Weak |
| Contrasts | 1 |
| Multi-slice mode | Sequential |
| Bandwidth | 540 Hz/Px |

### Sequence - Part 2

| | |
|---|---|
| RF pulse type | Fast |
| Gradient mode | Normal |
| Excitation | Non-sel. |
| RF spoiling | On |

### Sequence - Assistant

| | |
|---|---|
| Mode | Off |

**\\USER\WBIC Protocols Ready\Protocol 569\P00569_Epilepsy_20230628\flair_ns_tse_vfl_0.8iso_sag_UP**

TA: 6:54 PM: FIX Voxel size: 0.8×0.8×0.8 mmPAT: 6 Rel. SNR: 1.00 : spcir

### Properties

| | |
|---|---|
| Prio recon | Off |
| Load images to viewer | On |
| Inline movie | Off |
| Auto store images | On |
| Load images to stamp segments | Off |
| Load images to graphic segments | Off |
| Auto open inline display | Off |
| Auto close inline display | Off |
| Start measurement without further preparation | Off |
| Wait for user to start | Off |
| Start measurements | Single measurement |

### Routine

| | |
|---|---|
| Slab group | 1 |
| Slabs | 1 |
| Position | Isocenter |
| Orientation | Sagittal |
| Phase enc. dir. | A >> P |
| AutoAlign | --- |
| Phase oversampling | 0 % |
| Slice oversampling | 0.0 % |
| Slices per slab | 192 |
| FoV read | 230 mm |
| FoV phase | 100.0 % |
| Slice thickness | 0.80 mm |
| TR | 9000 ms |
| TE | 326 ms |
| Averages | 1.0 |
| Concatenations | 1 |
| Filter | Raw filter |
| Coil elements | AC |

### Contrast - Common

| | |
|---|---|
| TR | 9000 ms |
| TE | 326 ms |
| MTC | Off |
| Magn. preparation | Non-sel. T2-IR |
| TI | 2300 ms |
| T2prep duration | 100 ms |
| Fat suppr. | None |
| Blood suppr. | Off |
| Restore magn. | Off |

### Contrast - Dynamic

| | |
|---|---|
| Averages | 1.0 |
| Reconstruction | Magnitude |
| Measurements | 1 |
| Multiple series | Each measurement |

### Resolution - Common

| | |
|---|---|
| FoV read | 230 mm |
| FoV phase | 100.0 % |
| Slice thickness | 0.80 mm |
| Base resolution | 288 |
| Phase resolution | 100 % |
| Slice resolution | 100 % |
| Phase partial Fourier | Allowed |
| Slice partial Fourier | Off |

### Resolution - Common

| | |
|---|---|
| Interpolation | Off |

### Resolution - iPAT

| | |
|---|---|
| PAT mode | CAIPIRINHA |
| Accel. factor PE | 3 |
| Ref. lines PE | 32 |
| Accel. factor 3D | 2 |
| Ref. lines 3D | 24 |
| Reordering Shift 3D | 1 |
| Reference scan mode | Integrated |

### Resolution - Filter Image

| | |
|---|---|
| Image Filter | Off |
| Distortion Corr. | Off |
| Prescan Normalize | Off |
| Normalize | Off |
| B1 filter | Off |

### Resolution - Filter Rawdata

| | |
|---|---|
| Raw filter | On |
| Elliptical filter | Off |

### Geometry - Common

| | |
|---|---|
| Slab group | 1 |
| Slabs | 1 |
| Position | Isocenter |
| Orientation | Sagittal |
| Phase enc. dir. | A >> P |
| Slice oversampling | 0.0 % |
| Slices per slab | 192 |
| FoV read | 230 mm |
| FoV phase | 100.0 % |
| Slice thickness | 0.80 mm |
| TR | 9000 ms |
| Series | Ascending |
| Concatenations | 1 |

### Geometry - AutoAlign

| | |
|---|---|
| Slab group | 1 |
| Position | Isocenter |
| Orientation | Sagittal |
| Phase enc. dir. | A >> P |
| AutoAlign | --- |
| Initial Position | Isocenter |
| L | 0.0 mm |
| P | 0.0 mm |
| H | 0.0 mm |
| Initial Rotation | 0.00 deg |
| Initial Orientation | Sagittal |

### Geometry - Saturation

| | |
|---|---|
| Fat suppr. | None |
| Restore magn. | Off |
| Special sat. | None |

### Geometry - Navigator

### Geometry - Tim Planning Suite

| | |
|---|---|
| Set-n-Go Protocol | Off |
| Table position | H |
| Table position | 0 mm |
| Inline Composing | Off |

### System - Miscellaneous

| | |
|---|---|
| Positioning mode | FIX |
| Table position | H |
| Table position | 0 mm |
| MSMA | S - C - T |
| Sagittal | R >> L |
| Coronal | A >> P |
| Transversal | F >> H |
| Coil Combine Mode | Adaptive Combine |
| Save uncombined | Off |
| Matrix Optimization | Off |
| AutoAlign | --- |
| Coil Select Mode | Default |

### System - Adjustments

| | |
|---|---|
| B0 Shim mode | Brain |
| B1 Shim mode | TrueForm |
| Confirm freq. adjustment | Off |
| Assume Dominant Fat | Off |
| Assume Silicone | Off |
| Adjustment Tolerance | Auto |

### System - Adjust Volume

| | |
|---|---|
| ! Position | R0.9 P9.8 H15.0 mm |
| ! Orientation | T > C-16.4 > S-1.3 |
| ! Rotation | 2.64 deg |
| ! A >> P | 163 mm |
| ! R >> L | 160 mm |
| ! F >> H | 111 mm |
| Reset | Off |

### System - pTx Volumes

| | |
|---|---|
| B1 Shim mode | TrueForm |
| Excitation | Non-sel. |

### System - Tx/Rx

| | |
|---|---|
| Frequency 1H | 297.231131 MHz |
| Correction factor | 1 |
| Gain | High |
| Img. Scale Cor. | 1.000 |
| Reset | Off |
| ? Ref. amplitude 1H | 0.000 V |

### Physio - Signal1

| | |
|---|---|
| 1st Signal/Mode | None |
| Trigger delay | 0 ms |
| TR | 9000 ms |
| Concatenations | 1 |

### Physio - Cardiac

| | |
|---|---|
| Magn. preparation | Non-sel. T2-IR |
| TI | 2300 ms |
| T2prep duration | 100 ms |
| Fat suppr. | None |
| Dark blood | Off |
| FoV read | 230 mm |
| FoV phase | 100.0 % |
| Phase resolution | 100 % |

### Physio - PACE

| | |
|---|---|
| Resp. control | Off |
| Concatenations | 1 |

### Inline - Common

| | |
|---|---|
| Subtract | Off |
| Measurements | 1 |
| StdDev | Off |
| Save original images | On |

### Inline - MIP

| | |
|---|---|
| MIP-Sag | Off |
| MIP-Cor | Off |
| MIP-Tra | Off |
| MIP-Time | Off |
| Save original images | On |

### Inline - Composing

| | |
|---|---|
| Inline Composing | Off |
| Distortion Corr. | Off |

### Sequence - Part 1

| | |
|---|---|
| Introduction | Off |
| Dimension | 3D |
| Elliptical scanning | Off |
| Reordering | Linear |
| Flow comp. | No |
| Echo spacing | 5.48 ms |
| Bandwidth | 294 Hz/Px |

### Sequence - Part 2

| | |
|---|---|
| Echo train duration | 915 ms |
| Gradient mode | Fast |
| Excitation | Non-sel. |
| Flip angle mode | T2 var |
| Turbo factor | 218 |

### Sequence - pTX Pulses

| | |
|---|---|
| Universal Pulse | 1 |
| Pulse type | Excitation |
| Trajectory | External |

### Sequence - Assistant

| | |
|---|---|
| Allowed delay | 0 s |

**\\USER\WBIC Protocols Ready\Protocol 569\P00569_Epilepsy_20230628\Highresolution_TSE_PAT3_100_CTR1**

TA: 6:32 PM: REF Voxel size: 0.5×0.5×1.0 mmPAT: 3 Rel. SNR: 1.00 : qtse

### Properties

| | |
|---|---|
| Prio recon | Off |
| Load images to viewer | On |
| Inline movie | Off |
| Auto store images | On |
| Load images to stamp segments | Off |
| Load images to graphic segments | Off |
| Auto open inline display | Off |
| Auto close inline display | Off |
| Start measurement without further preparation | Off |
| Wait for user to start | Off |
| Start measurements | Single measurement |

### Routine

| | |
|---|---|
| Slice group | 1 |
| Slices | 120 |
| Dist. factor | 10 % |
| Position | L0.0 P3.2 F8.5 mm |
| Orientation | C > T-14.7 > S0.1 |
| Phase enc. dir. | R >> L |
| AutoAlign | --- |
| Phase oversampling | 0 % |
| FoV read | 244 mm |
| FoV phase | 100.0 % |
| Slice thickness | 1.0 mm |
| TR | 8870.0 ms |
| TE | 77 ms |
| Averages | 1 |
| Concatenations | 2 |
| Filter | Raw filter |
| Coil elements | AC |

### Contrast - Common

| | |
|---|---|
| TR | 8870.0 ms |
| TE | 77 ms |
| TD | 0.0 ms |
| MTC | Off |
| Magn. preparation | None |
| Flip angle | 60 deg |
| Fat suppr. | None |
| Water suppr. | None |
| Restore magn. | Off |

### Contrast - Dynamic

| | |
|---|---|
| Averages | 1 |
| Averaging mode | Short term |
| Reconstruction | Magnitude |
| Measurements | 1 |
| Multiple series | Each measurement |

### Resolution - Common

| | |
|---|---|
| FoV read | 244 mm |
| FoV phase | 100.0 % |
| Slice thickness | 1.0 mm |
| Base resolution | 512 |
| Phase resolution | 100 % |
| Phase partial Fourier | Off |
| Trajectory | Cartesian |
| Interpolation | Off |

### Resolution - iPAT

| | |
|---|---|
| PAT mode | GRAPPA |
| Accel. factor PE | 3 |
| Ref. lines PE | 27 |
| Reference scan mode | Integrated |

### Resolution - Filter Image

| | |
|---|---|
| Image Filter | Off |
| Distortion Corr. | Off |
| Prescan Normalize | Off |
| Normalize | Off |
| B1 filter | Off |

### Resolution - Filter Rawdata

| | |
|---|---|
| Raw filter | On |
| Elliptical filter | Off |

### Geometry - Common

| | |
|---|---|
| Slice group | 1 |
| Slices | 120 |
| Dist. factor | 10 % |
| Position | L0.0 P3.2 F8.5 mm |
| Orientation | C > T-14.7 > S0.1 |
| Phase enc. dir. | R >> L |
| FoV read | 244 mm |
| FoV phase | 100.0 % |
| Slice thickness | 1.0 mm |
| TR | 8870.0 ms |
| Multi-slice mode | Interleaved |
| Series | Interleaved |
| Concatenations | 2 |

### Geometry - AutoAlign

| | |
|---|---|
| Slice group | 1 |
| Position | L0.0 P3.2 F8.5 mm |
| Orientation | C > T-14.7 > S0.1 |
| Phase enc. dir. | R >> L |
| AutoAlign | --- |
| Initial Position | L0.0 P3.2 F8.5 |
| R | 0.0 mm |
| P | 3.2 mm |
| F | 8.5 mm |
| Initial Rotation | -1.30 deg |
| Initial Orientation | C > T |
| C > T | -14.7 |
| > S | 0.1 |

### Geometry - Saturation

| | |
|---|---|
| Fat suppr. | None |
| Water suppr. | None |
| Restore magn. | Off |
| Special sat. | None |

### Geometry - Navigator

### Geometry - Tim Planning Suite

| | |
|---|---|
| Set-n-Go Protocol | Off |
| Table position | H |
| Table position | 0 mm |

#### Geometry - Tim Planning Suite

| | |
|---|---|
| Inline Composing | Off |

#### Geometry - Tim CT

| | |
|---|---|
| Tim CT mode | Off |
| Slices | 120 |
| Slice thickness | 1.0 mm |
| Dist. factor | 10 % |
| FoV read | 244 mm |
| FoV phase | 100.0 % |

#### System - Miscellaneous

| | |
|---|---|
| Positioning mode | REF |
| Table position | F |
| Table position | 0 mm |
| MSMA | S - C - T |
| Sagittal | R >> L |
| Coronal | A >> P |
| Transversal | F >> H |
| Coil Combine Mode | Sum of Squares |
| Save uncombined | Off |
| Matrix Optimization | Off |
| AutoAlign | --- |
| Coil Select Mode | Default |

#### System - Adjustments

| | |
|---|---|
| B0 Shim mode | Brain |
| B1 Shim mode | TrueForm |
| Confirm freq. adjustment | Off |
| Assume Dominant Fat | Off |
| Assume Silicone | Off |
| Adjustment Tolerance | Auto |

#### System - Adjust Volume

| | |
|---|---|
| ! Position | R0.9 P9.8 H15.0 mm |
| ! Orientation | T > C-16.4 > S-1.3 |
| ! Rotation | 2.64 deg |
| ! A >> P | 163 mm |
| ! R >> L | 160 mm |
| ! F >> H | 111 mm |
| Reset | Off |

#### System - pTx Volumes

| | |
|---|---|
| B1 Shim mode | TrueForm |

#### System - Tx/Rx

| | |
|---|---|
| Frequency 1H | 297.231131 MHz |
| Correction factor | 1 |
| Gain | High |
| Img. Scale Cor. | 1.000 |
| Reset | Off |
| ? Ref. amplitude 1H | 0.000 V |

#### Physio - Signal1

| | |
|---|---|
| 1st Signal/Mode | None |
| TR | 8870.0 ms |
| Concatenations | 2 |

#### Physio - Cardiac

| | |
|---|---|
| Magn. preparation | None |
| Fat suppr. | None |
| Dark blood | Off |
| FoV read | 244 mm |
| FoV phase | 100.0 % |

#### Physio - Cardiac

| | |
|---|---|
| Phase resolution | 100 % |
| Trajectory | Cartesian |

#### Physio - PACE

| | |
|---|---|
| Resp. control | Off |
| Concatenations | 2 |

#### Inline - Common

| | |
|---|---|
| Subtract | Off |
| Measurements | 1 |
| StdDev | Off |
| Save original images | On |

#### Inline - MIP

| | |
|---|---|
| MIP-Sag | Off |
| MIP-Cor | Off |
| MIP-Tra | Off |
| MIP-Time | Off |
| Save original images | On |

#### Inline - Composing

| | |
|---|---|
| Inline Composing | Off |
| Distortion Corr. | Off |

#### Sequence - Part 1

| | |
|---|---|
| Introduction | On |
| Dimension | 2D |
| Compensate T2 decay | Off |
| Reduce Motion Sens. | Off |
| Contrasts | 1 |
| Flow comp. | No |
| Optimization | In phase |
| Multi-slice mode | Interleaved |
| Free echo spacing | On |
| Echo spacing | 15.3 ms |
| Bandwidth | 178 Hz/Px |

#### Sequence - Part 2

| | |
|---|---|
| Define | Turbo factor |
| Echo trains per slice | 21 |
| Phase correction | Automatic |
| Acoustic noise reduction | Active |
| RF pulse type | Low SAR |
| Gradient mode | Normal |
| Hyperecho | On |
| WARP | Off |
| Red. EC sensitivity | Off |
| Turbo factor | 9 |

#### Sequence - Assistant

| | |
|---|---|
| Mode | Off |
| Allowed delay | 0 s |

**\\USER\WBIC Protocols Ready\Protocol 569\P00569_Epilepsy_20230628\t2_ns_tse_vfl_0.8iso_sag_UP**

TA: 8:00 PM: FIX Voxel size: 0.8×0.8×0.8 mmPAT: 9 Rel. SNR: 1.00 : spc

### Properties

| | |
|---|---|
| Prio recon | Off |
| Load images to viewer | On |
| Inline movie | Off |
| Auto store images | On |
| Load images to stamp segments | Off |
| Load images to graphic segments | Off |
| Auto open inline display | Off |
| Auto close inline display | Off |
| Start measurement without further preparation | Off |
| Wait for user to start | Off |
| Start measurements | Single measurement |

### Routine

| | |
|---|---|
| Slab group | 1 |
| Slabs | 1 |
| Position | Isocenter |
| Orientation | Sagittal |
| Phase enc. dir. | A >> P |
| AutoAlign | --- |
| Phase oversampling | 0 % |
| Slice oversampling | 0.0 % |
| Slices per slab | 240 |
| FoV read | 230 mm |
| FoV phase | 100.0 % |
| Slice thickness | 0.80 mm |
| TR | 12000 ms |
| TE | 384 ms |
| Averages | 1.0 |
| Concatenations | 1 |
| Filter | Raw filter |
| Coil elements | AC |

### Contrast - Common

| | |
|---|---|
| TR | 12000 ms |
| TE | 384 ms |
| MTC | Off |
| Magn. preparation | None |
| Fat suppr. | None |
| Blood suppr. | Off |
| Restore magn. | Off |

### Contrast - Dynamic

| | |
|---|---|
| Averages | 1.0 |
| Reconstruction | Magnitude |
| Measurements | 1 |
| Multiple series | Each measurement |

### Resolution - Common

| | |
|---|---|
| FoV read | 230 mm |
| FoV phase | 100.0 % |
| Slice thickness | 0.80 mm |
| Base resolution | 288 |
| Phase resolution | 100 % |
| Slice resolution | 100 % |
| Phase partial Fourier | Allowed |
| Slice partial Fourier | Off |
| Interpolation | Off |

### Resolution - iPAT

| | |
|---|---|
| PAT mode | CAIPIRINHA |
| Accel. factor PE | 3 |
| Ref. lines PE | 32 |
| Accel. factor 3D | 3 |
| Ref. lines 3D | 24 |
| Reordering Shift 3D | 1 |
| Reference scan mode | Integrated |

### Resolution - Filter Image

| | |
|---|---|
| Image Filter | Off |
| Distortion Corr. | Off |
| Prescan Normalize | Off |
| Normalize | Off |
| B1 filter | Off |

### Resolution - Filter Rawdata

| | |
|---|---|
| Raw filter | On |
| Elliptical filter | Off |

### Geometry - Common

| | |
|---|---|
| Slab group | 1 |
| Slabs | 1 |
| Position | Isocenter |
| Orientation | Sagittal |
| Phase enc. dir. | A >> P |
| Slice oversampling | 0.0 % |
| Slices per slab | 240 |
| FoV read | 230 mm |
| FoV phase | 100.0 % |
| Slice thickness | 0.80 mm |
| TR | 12000 ms |
| Series | Ascending |
| Concatenations | 1 |

### Geometry - AutoAlign

| | |
|---|---|
| Slab group | 1 |
| Position | Isocenter |
| Orientation | Sagittal |
| Phase enc. dir. | A >> P |
| AutoAlign | --- |
| Initial Position | Isocenter |
| L | 0.0 mm |
| P | 0.0 mm |
| H | 0.0 mm |
| Initial Rotation | 0.00 deg |
| Initial Orientation | Sagittal |

### Geometry - Saturation

| | |
|---|---|
| Fat suppr. | None |
| Restore magn. | Off |
| Special sat. | None |

### Geometry - Navigator

### Geometry - Tim Planning Suite

| | |
|---|---|
| Set-n-Go Protocol | Off |
| Table position | H |
| Table position | 0 mm |

#### Geometry - Tim Planning Suite

| | |
|---|---|
| Inline Composing | Off |

#### System - Miscellaneous

| | |
|---|---|
| Positioning mode | FIX |
| Table position | H |
| Table position | 0 mm |
| MSMA | S - C - T |
| Sagittal | R >> L |
| Coronal | A >> P |
| Transversal | F >> H |
| Coil Combine Mode | Adaptive Combine |
| Save uncombined | Off |
| Matrix Optimization | Off |
| AutoAlign | --- |
| Coil Select Mode | Default |

#### System - Adjustments

| | |
|---|---|
| B0 Shim mode | Brain |
| B1 Shim mode | TrueForm |
| Confirm freq. adjustment | Off |
| Assume Dominant Fat | Off |
| Assume Silicone | Off |
| Adjustment Tolerance | Auto |

#### System - Adjust Volume

| | |
|---|---|
| ! Position | R0.9 P9.8 H15.0 mm |
| ! Orientation | T > C-16.4 > S-1.3 |
| ! Rotation | 2.64 deg |
| ! A >> P | 163 mm |
| ! R >> L | 160 mm |
| ! F >> H | 111 mm |
| Reset | Off |

#### System - pTx Volumes

| | |
|---|---|
| B1 Shim mode | TrueForm |
| Excitation | Non-sel. |

#### System - Tx/Rx

| | |
|---|---|
| Frequency 1H | 297.231131 MHz |
| Correction factor | 1 |
| Gain | High |
| Img. Scale Cor. | 1.000 |
| Reset | Off |
| ? Ref. amplitude 1H | 0.000 V |

#### Physio - Signal1

| | |
|---|---|
| 1st Signal/Mode | None |
| Trigger delay | 0 ms |
| TR | 12000 ms |
| Concatenations | 1 |

#### Physio - Cardiac

| | |
|---|---|
| Magn. preparation | None |
| Fat suppr. | None |
| Dark blood | Off |
| FoV read | 230 mm |
| FoV phase | 100.0 % |
| Phase resolution | 100 % |

#### Physio - PACE

| | |
|---|---|
| Resp. control | Off |
| Concatenations | 1 |

#### Inline - Common

| | |
|---|---|
| Subtract | Off |
| Measurements | 1 |
| StdDev | Off |
| Save original images | On |

#### Inline - MIP

| | |
|---|---|
| MIP-Sag | Off |
| MIP-Cor | Off |
| MIP-Tra | Off |
| MIP-Time | Off |
| Save original images | On |

#### Inline - Composing

| | |
|---|---|
| Inline Composing | Off |
| Distortion Corr. | Off |

#### Sequence - Part 1

| | |
|---|---|
| Introduction | Off |
| Dimension | 3D |
| Elliptical scanning | Off |
| Reordering | Linear |
| Flow comp. | No |
| Echo spacing | 3.52 ms |
| Bandwidth | 694 Hz/Px |

#### Sequence - Part 2

| | |
|---|---|
| Echo train duration | 764 ms |
| Gradient mode | Fast |
| Excitation | Non-sel. |
| Flip angle mode | T2 var |
| Turbo factor | 218 |

#### Sequence - pTX Pulses

| | |
|---|---|
| Universal Pulse | 1 |
| Pulse type | Excitation |
| Trajectory | External |

#### Sequence - Assistant

| | |
|---|---|
| Allowed delay | 0 s |

**\\USER\WBIC Protocols Ready\Protocol 569\P00569_Epilepsy_20230628\T2star_0.8iso_GRAPPA_2_2**

TA: 7:17 PM: FIX Voxel size: 0.8×0.8×0.8 mmPAT: 4 Rel. SNR: 1.00 : fl_r

### Properties

| | |
|---|---|
| Prio recon | Off |
| Load images to viewer | On |
| Inline movie | Off |
| Auto store images | On |
| Load images to stamp segments | Off |
| Load images to graphic segments | Off |
| Auto open inline display | Off |
| Auto close inline display | Off |
| Start measurement without further preparation | Off |
| Wait for user to start | Off |
| Start measurements | Single measurement |

### Routine

| | |
|---|---|
| Slab group | 1 |
| Slabs | 1 |
| Dist. factor | 20 % |
| Position | Isocenter |
| Orientation | Sagittal |
| Phase enc. dir. | A >> P |
| AutoAlign | --- |
| Phase oversampling | 0 % |
| Slice oversampling | 0.0 % |
| Slices per slab | 192 |
| FoV read | 224 mm |
| FoV phase | 100.0 % |
| Slice thickness | 0.80 mm |
| TR | 31.0 ms |
| TE | 20.00 ms |
| Averages | 1 |
| Concatenations | 1 |
| Filter | None |
| Coil elements | AC |

### Contrast - Common

| | |
|---|---|
| TR | 31.0 ms |
| TE | 20.00 ms |
| MTC | Off |
| Magn. preparation | None |
| Flip angle | 15 deg |
| Fat suppr. | None |
| Water suppr. | None |
| SWI | Off |

### Contrast - Dynamic

| | |
|---|---|
| Averages | 1 |
| Averaging mode | Short term |
| Reconstruction | Magn./Phase |
| Measurements | 1 |
| Multiple series | Each measurement |

### Resolution - Common

| | |
|---|---|
| FoV read | 224 mm |
| FoV phase | 100.0 % |
| Slice thickness | 0.80 mm |
| Base resolution | 280 |
| Phase resolution | 100 % |
| Slice resolution | 100 % |
| Phase partial Fourier | Off |

### Resolution - Common

| | |
|---|---|
| Slice partial Fourier | Off |
| Interpolation | Off |

### Resolution - iPAT

| | |
|---|---|
| PAT mode | GRAPPA |
| Accel. factor PE | 2 |
| Ref. lines PE | 40 |
| Accel. factor 3D | 2 |
| Ref. lines 3D | 24 |
| Reference scan mode | Integrated |

### Resolution - Filter Image

| | |
|---|---|
| Image Filter | Off |
| Distortion Corr. | Off |
| Prescan Normalize | Off |
| Normalize | Off |
| B1 filter | Off |

### Resolution - Filter Rawdata

| | |
|---|---|
| Raw filter | Off |
| Elliptical filter | Off |

### Geometry - Common

| | |
|---|---|
| Slab group | 1 |
| Slabs | 1 |
| Dist. factor | 20 % |
| Position | Isocenter |
| Orientation | Sagittal |
| Phase enc. dir. | A >> P |
| Slice oversampling | 0.0 % |
| Slices per slab | 192 |
| FoV read | 224 mm |
| FoV phase | 100.0 % |
| Slice thickness | 0.80 mm |
| TR | 31.0 ms |
| Multi-slice mode | Interleaved |
| Series | Interleaved |
| Concatenations | 1 |

### Geometry - AutoAlign

| | |
|---|---|
| Slab group | 1 |
| Position | Isocenter |
| Orientation | Sagittal |
| Phase enc. dir. | A >> P |
| AutoAlign | --- |
| Initial Position | Isocenter |
| L | 0.0 mm |
| P | 0.0 mm |
| H | 0.0 mm |
| Initial Rotation | 0.00 deg |
| Initial Orientation | Sagittal |

### Geometry - Saturation

| | |
|---|---|
| Saturation mode | Standard |
| Fat suppr. | None |
| Water suppr. | None |
| Special sat. | None |

#### Geometry - Tim Planning Suite

| | |
|---|---|
| Set-n-Go Protocol | Off |
| Table position | H |
| Table position | 0 mm |
| Inline Composing | Off |

#### Geometry - Tim CT

| | |
|---|---|
| Tim CT mode | Off |
| Slabs | 1 |
| Slices per slab | 192 |
| Slice thickness | 0.80 mm |
| Dist. factor | 20 % |
| FoV read | 224 mm |
| FoV phase | 100.0 % |
| Segments | 1 |

#### System - Miscellaneous

| | |
|---|---|
| Positioning mode | FIX |
| Table position | F |
| Table position | 0 mm |
| MSMA | S - C - T |
| Sagittal | R >> L |
| Coronal | A >> P |
| Transversal | F >> H |
| Coil Combine Mode | Sum of Squares |
| Save uncombined | Off |
| Matrix Optimization | Off |
| AutoAlign | --- |
| Coil Select Mode | Default |

#### System - Adjustments

| | |
|---|---|
| B0 Shim mode | Brain |
| B1 Shim mode | TrueForm |
| Confirm freq. adjustment | Off |
| Assume Dominant Fat | Off |
| Assume Silicone | Off |
| Adjustment Tolerance | None |

#### System - Adjust Volume

| | |
|---|---|
| ! Position | R0.9 P9.8 H15.0 mm |
| ! Orientation | T > C-16.4 > S-1.3 |
| ! Rotation | 2.64 deg |
| ! A >> P | 163 mm |
| ! R >> L | 160 mm |
| ! F >> H | 111 mm |
| Reset | Off |

#### System - pTx Volumes

| | |
|---|---|
| B1 Shim mode | TrueForm |
| Excitation | Slab-sel. |

#### System - Tx/Rx

| | |
|---|---|
| Frequency 1H | 297.231131 MHz |
| Correction factor | 1 |
| Gain | High |
| Img. Scale Cor. | 1.000 |
| Reset | Off |
| ? Ref. amplitude 1H | 0.000 V |

#### Physio - Signal1

| | |
|---|---|
| 1st Signal/Mode | None |
| TR | 31.0 ms |
| Concatenations | 1 |
| Segments | 1 |

#### Physio - Cardiac

| | |
|---|---|
| Tagging | None |
| Magn. preparation | None |
| Fat suppr. | None |
| Dark blood | Off |
| FoV read | 224 mm |
| FoV phase | 100.0 % |
| Phase resolution | 100 % |

#### Physio - PACE

| | |
|---|---|
| Resp. control | Off |
| Concatenations | 1 |

#### Inline - Common

| | |
|---|---|
| Subtract | Off |
| Measurements | 1 |
| StdDev | Off |
| Liver registration | Off |
| Save original images | On |

#### Inline - MIP

| | |
|---|---|
| MIP-Sag | Off |
| MIP-Cor | Off |
| MIP-Tra | Off |
| MIP-Time | Off |
| Save original images | On |

#### Inline - Soft Tissue

| | |
|---|---|
| Wash - In | Off |
| Wash - Out | Off |
| TTP | Off |
| PEI | Off |
| MIP - time | Off |
| Measurements | 1 |

#### Inline - Composing

| | |
|---|---|
| Inline Composing | Off |
| Distortion Corr. | Off |

#### Inline - MapIt

| | |
|---|---|
| Save original images | On |
| MapIt | None |
| Flip angle | 15 deg |
| Measurements | 1 |
| Contrasts | 1 |
| TR | 31.0 ms |
| TE | 20.00 ms |

#### Sequence - Part 1

| | |
|---|---|
| Introduction | On |
| Dimension | 3D |
| Elliptical scanning | Off |
| Phase stabilisation | Off |
| Asymmetric echo | Off |
| Contrasts | 1 |
| Flow comp. | Slice/Read |
| Multi-slice mode | Interleaved |
| Bandwidth | 80 Hz/Px |

#### Sequence - Part 2

| | |
|---|---|
| Segments | 1 |
| Acoustic noise reduction | None |
| RF pulse type | Normal |
| Gradient mode | Fast |

#### Sequence - Part 2

| | |
|---|---|
| Excitation | Slab-sel. |
| RF spoiling | On |

#### Sequence - Assistant

| | |
|---|---|
| Mode | Off |

**\\USER\WBIC Protocols Ready\Protocol 569\P00569_Epilepsy_20230628\dark-fluid_spcR_sag_p6_0.8**

TA: 7:23 PM: FIX Voxel size: 0.8×0.8×0.8 mmPAT: 6 Rel. SNR: 1.00 : spcir

### Properties

| | |
|---|---|
| Prio recon | Off |
| Load images to viewer | On |
| Inline movie | Off |
| Auto store images | On |
| Load images to stamp segments | Off |
| Load images to graphic segments | Off |
| Auto open inline display | Off |
| Auto close inline display | Off |
| Start measurement without further preparation | Off |
| Wait for user to start | Off |
| Start measurements | Single measurement |

### Routine

| | |
|---|---|
| Slab group | 1 |
| Slabs | 1 |
| Position | Isocenter |
| Orientation | Sagittal |
| Phase enc. dir. | A >> P |
| AutoAlign | --- |
| Phase oversampling | 0 % |
| Slice oversampling | 8.3 % |
| Slices per slab | 192 |
| FoV read | 230 mm |
| FoV phase | 100.0 % |
| Slice thickness | 0.80 mm |
| TR | 9000 ms |
| TE | 269 ms |
| Averages | 1.0 |
| Concatenations | 1 |
| Filter | B1 filter |
| Coil elements | AC |

### Contrast - Common

| | |
|---|---|
| TR | 9000 ms |
| TE | 269 ms |
| MTC | Off |
| Magn. preparation | Non-sel. T2-IR |
| TI 1 | 2600 ms |
| Fat suppr. | None |
| Blood suppr. | Off |
| Restore magn. | Off |

### Contrast - Dynamic

| | |
|---|---|
| Averages | 1.0 |
| Reconstruction | Magnitude |
| Measurements | 1 |
| Multiple series | Each measurement |

### Resolution - Common

| | |
|---|---|
| FoV read | 230 mm |
| FoV phase | 100.0 % |
| Slice thickness | 0.80 mm |
| Base resolution | 288 |
| Phase resolution | 100 % |
| Slice resolution | 100 % |
| Phase partial Fourier | Allowed |
| Slice partial Fourier | Off |
| Interpolation | Off |

### Resolution - iPAT

| | |
|---|---|
| PAT mode | CAIPIRINHA |
| Accel. factor PE | 3 |
| Ref. lines PE | 32 |
| Accel. factor 3D | 2 |
| Ref. lines 3D | 24 |
| Reordering Shift 3D | 1 |
| Reference scan mode | Integrated |

### Resolution - Filter Image

| | |
|---|---|
| Image Filter | Off |
| Distortion Corr. | Off |
| Prescan Normalize | Off |
| Normalize | Off |
| B1 filter | On |
| Unfiltered images | Off |

### Resolution - Filter Rawdata

| | |
|---|---|
| Raw filter | Off |
| Elliptical filter | Off |

### Geometry - Common

| | |
|---|---|
| Slab group | 1 |
| Slabs | 1 |
| Position | Isocenter |
| Orientation | Sagittal |
| Phase enc. dir. | A >> P |
| Slice oversampling | 8.3 % |
| Slices per slab | 192 |
| FoV read | 230 mm |
| FoV phase | 100.0 % |
| Slice thickness | 0.80 mm |
| TR | 9000 ms |
| Series | Ascending |
| Concatenations | 1 |

### Geometry - AutoAlign

| | |
|---|---|
| Slab group | 1 |
| Position | Isocenter |
| Orientation | Sagittal |
| Phase enc. dir. | A >> P |
| AutoAlign | --- |
| Initial Position | Isocenter |
| L | 0.0 mm |
| P | 0.0 mm |
| H | 0.0 mm |
| Initial Rotation | 0.00 deg |
| Initial Orientation | Sagittal |

### Geometry - Saturation

| | |
|---|---|
| Fat suppr. | None |
| Restore magn. | Off |
| Special sat. | None |

### Geometry - Navigator

### Geometry - Tim Planning Suite

| | |
|---|---|
| Set-n-Go Protocol | Off |
| Table position | H |

### Geometry - Tim Planning Suite

| | |
|---|---|
| Table position | 0 mm |
| Inline Composing | Off |

### System - Miscellaneous

| | |
|---|---|
| Positioning mode | FIX |
| Table position | H |
| Table position | 0 mm |
| MSMA | S - C - T |
| Sagittal | R >> L |
| Coronal | A >> P |
| Transversal | F >> H |
| Coil Combine Mode | Adaptive Combine |
| Save uncombined | Off |
| Matrix Optimization | Off |
| AutoAlign | --- |
| Coil Select Mode | Default |

### System - Adjustments

| | |
|---|---|
| B0 Shim mode | Brain |
| B1 Shim mode | TrueForm |
| Confirm freq. adjustment | Off |
| Assume Dominant Fat | Off |
| Assume Silicone | Off |
| Adjustment Tolerance | Auto |

### System - Adjust Volume

| | |
|---|---|
| ! Position | R0.9 P9.8 H15.0 mm |
| ! Orientation | T > C-16.4 > S-1.3 |
| ! Rotation | 2.64 deg |
| ! A >> P | 163 mm |
| ! R >> L | 160 mm |
| ! F >> H | 111 mm |
| Reset | Off |

### System - pTx Volumes

| | |
|---|---|
| B1 Shim mode | TrueForm |
| Excitation | Non-sel. |

### System - Tx/Rx

| | |
|---|---|
| Frequency 1H | 297.231131 MHz |
| Correction factor | 1 |
| Gain | High |
| Img. Scale Cor. | 5.000 |
| Reset | Off |
| ? Ref. amplitude 1H | 0.000 V |

### Physio - Signal1

| | |
|---|---|
| 1st Signal/Mode | None |
| Trigger delay | 0 ms |
| TR | 9000 ms |
| Concatenations | 1 |

### Physio - Cardiac

| | |
|---|---|
| Magn. preparation | Non-sel. T2-IR |
| TI 1 | 2600 ms |
| Fat suppr. | None |
| Dark blood | Off |
| FoV read | 230 mm |
| FoV phase | 100.0 % |
| Phase resolution | 100 % |

### Physio - PACE

| | |
|---|---|
| Resp. control | Off |

### Physio - PACE

| | |
|---|---|
| Concatenations | 1 |

### Inline - Common

| | |
|---|---|
| Subtract | Off |
| Measurements | 1 |
| StdDev | Off |
| Save original images | On |

### Inline - MIP

| | |
|---|---|
| MIP-Sag | Off |
| MIP-Cor | Off |
| MIP-Tra | Off |
| MIP-Time | Off |
| Save original images | On |

### Inline - Composing

| | |
|---|---|
| Inline Composing | Off |
| Distortion Corr. | Off |

### Sequence - Part 1

| | |
|---|---|
| Introduction | On |
| Dimension | 3D |
| Elliptical scanning | Off |
| Reordering | Linear |
| Flow comp. | No |
| Echo spacing | 3.32 ms |
| Adiabatic-mode | Off |
| Bandwidth | 723 Hz/Px |

### Sequence - Part 2

| | |
|---|---|
| Echo train duration | 631 ms |
| RF pulse type | Normal |
| Gradient mode | Fast |
| Excitation | Non-sel. |
| Flip angle mode | T2 var |
| Turbo factor | 220 |

### Sequence - Assistant

| | |
|---|---|
| Allowed delay | 120 s |

**\\USER\WBIC Protocols Ready\Protocol 569\P00569_Epilepsy_20230628\mp2rage_sag_p3_0.8mm**

TA: 8:33 PM: REF Voxel size: 0.8×0.8×0.8 mmPAT: 3 Rel. SNR: 1.00 : tfl

### Properties

| | |
|---|---|
| Prio recon | Off |
| Load images to viewer | On |
| Inline movie | Off |
| Auto store images | On |
| Load images to stamp segments | Off |
| Load images to graphic segments | Off |
| Auto open inline display | Off |
| Auto close inline display | Off |
| Start measurement without further preparation | Off |
| Wait for user to start | Off |
| Start measurements | Single measurement |

### Routine

| | |
|---|---|
| Slab group | 1 |
| Slabs | 1 |
| Dist. factor | 50 % |
| Position | Isocenter |
| Orientation | Sagittal |
| Phase enc. dir. | A >> P |
| AutoAlign | --- |
| Phase oversampling | 0 % |
| Slice oversampling | 0.0 % |
| Slices per slab | 192 |
| FoV read | 240 mm |
| FoV phase | 100.0 % |
| Slice thickness | 0.80 mm |
| TR | 4300.0 ms |
| TE | 1.99 ms |
| Averages | 1 |
| Concatenations | 1 |
| Filter | None |
| Coil elements | AC |

### Contrast - Common

| | |
|---|---|
| TR | 4300.0 ms |
| TE | 1.99 ms |
| Magn. preparation | Non-sel. IR |
| TI 1 | 840 ms |
| TI 2 | 2370 ms |
| Flip angle 1 | 5.0 deg |
| Flip angle 2 | 6.0 deg |
| Fat suppr. | Water excit. fast |
| Water suppr. | None |

### Contrast - Dynamic

| | |
|---|---|
| Averages | 1 |
| Averaging mode | Long term |
| Reconstruction | Magnitude |
| Measurements | 1 |
| Multiple series | Each measurement |

### Resolution - Common

| | |
|---|---|
| FoV read | 240 mm |
| FoV phase | 100.0 % |
| Slice thickness | 0.80 mm |
| Base resolution | 288 |
| Phase resolution | 100 % |
| Slice resolution | 100 % |
| Phase partial Fourier | Off |

### Resolution - Common

| | |
|---|---|
| Slice partial Fourier | 6/8 |
| Interpolation | Off |

### Resolution - iPAT

| | |
|---|---|
| PAT mode | GRAPPA |
| Accel. factor PE | 3 |
| Ref. lines PE | 36 |
| Accel. factor 3D | 1 |
| Reference scan mode | Integrated |

### Resolution - Filter Image

| | |
|---|---|
| Image Filter | Off |
| Distortion Corr. | Off |
| Prescan Normalize | Off |
| Normalize | Off |
| B1 filter | Off |

### Resolution - Filter Rawdata

| | |
|---|---|
| Raw filter | Off |
| Elliptical filter | Off |

### Geometry - Common

| | |
|---|---|
| Slab group | 1 |
| Slabs | 1 |
| Dist. factor | 50 % |
| Position | Isocenter |
| Orientation | Sagittal |
| Phase enc. dir. | A >> P |
| Slice oversampling | 0.0 % |
| Slices per slab | 192 |
| FoV read | 240 mm |
| FoV phase | 100.0 % |
| Slice thickness | 0.80 mm |
| TR | 4300.0 ms |
| Multi-slice mode | Single shot |
| Series | Interleaved |
| Concatenations | 1 |

### Geometry - AutoAlign

| | |
|---|---|
| Slab group | 1 |
| Position | Isocenter |
| Orientation | Sagittal |
| Phase enc. dir. | A >> P |
| AutoAlign | --- |
| Initial Position | Isocenter |
| L | 0.0 mm |
| P | 0.0 mm |
| H | 0.0 mm |
| Initial Rotation | 21.00 deg |
| Initial Orientation | Sagittal |

### Geometry - Navigator

### Geometry - Tim Planning Suite

| | |
|---|---|
| Set-n-Go Protocol | Off |
| Table position | H |
| Table position | 0 mm |
| Inline Composing | Off |

### System - Miscellaneous

| | |
|---|---|
| Positioning mode | REF |
| Table position | H |
| Table position | 0 mm |
| MSMA | S - C - T |
| Sagittal | R >> L |
| Coronal | A >> P |
| Transversal | F >> H |
| Coil Combine Mode | Sum of Squares |
| Save uncombined | Off |
| Matrix Optimization | Off |
| AutoAlign | --- |
| Coil Select Mode | Default |

### System - Adjustments

| | |
|---|---|
| B0 Shim mode | Brain |
| B1 Shim mode | TrueForm |
| Confirm freq. adjustment | Off |
| Assume Dominant Fat | Off |
| Assume Silicone | Off |
| Adjustment Tolerance | Auto |

### System - Adjust Volume

| | |
|---|---|
| ! Position | R0.9 P9.8 H15.0 mm |
| ! Orientation | T > C-16.4 > S-1.3 |
| ! Rotation | 2.64 deg |
| ! A >> P | 163 mm |
| ! R >> L | 160 mm |
| ! F >> H | 111 mm |
| Reset | Off |

### System - pTx Volumes

| | |
|---|---|
| B1 Shim mode | TrueForm |
| Excitation | Non-sel. |

### System - Tx/Rx

| | |
|---|---|
| Frequency 1H | 297.231131 MHz |
| Correction factor | 1 |
| Gain | High |
| Img. Scale Cor. | 1.000 |
| Reset | Off |
| ? Ref. amplitude 1H | 0.000 V |

### Physio - Signal1

| | |
|---|---|
| 1st Signal/Mode | None |
| TR | 4300.0 ms |
| Concatenations | 1 |

### Physio - Cardiac

| | |
|---|---|
| Magn. preparation | Non-sel. IR |
| TI 1 | 840 ms |
| TI 2 | 2370 ms |
| Fat suppr. | Water excit. fast |
| Dark blood | Off |
| FoV read | 240 mm |
| FoV phase | 100.0 % |
| Phase resolution | 100 % |

### Physio - PACE

| | |
|---|---|
| Resp. control | Off |
| Concatenations | 1 |

### Inline - Common

| | |
|---|---|
| Subtract | Off |

### Inline - Common

| | |
|---|---|
| Measurements | 1 |
| StdDev | Off |
| Save original images | On |

### Inline - MIP

| | |
|---|---|
| MIP-Sag | Off |
| MIP-Cor | Off |
| MIP-Tra | Off |
| MIP-Time | Off |
| Save original images | On |

### Inline - Composing

| | |
|---|---|
| Inline Composing | Off |
| Distortion Corr. | Off |

### Inline - MapIt

| | |
|---|---|
| Save original images | On |
| MapIt | None |
| Flip angle 1 | 5.0 deg |
| Flip angle 2 | 6.0 deg |
| Measurements | 1 |
| TR | 4300.0 ms |
| TE | 1.99 ms |

### Sequence - Part 1

| | |
|---|---|
| Introduction | On |
| Dimension | 3D |
| Elliptical scanning | Off |
| Reordering | Linear |
| Asymmetric echo | Allowed |
| Flow comp. | No |
| Multi-slice mode | Single shot |
| Echo spacing | 6.3 ms |
| Bandwidth | 250 Hz/Px |

### Sequence - Part 2

| | |
|---|---|
| RF pulse type | Fast |
| Gradient mode | Fast* |
| Excitation | Non-sel. |
| RF spoiling | On |
| Incr. Gradient spoiling | Off |
| Turbo factor | 144 |

### Sequence - Assistant

| | |
|---|---|
| Mode | Off |

**\\USER\WBIC Protocols Ready\Protocol 569\P00569_Epilepsy_20230628\extra_ep2d_diff_PTX**

TA: 3:57 PM: FIX Voxel size: 1.3×1.3×1.3 mmPAT: 3 Rel. SNR: 1.00 : epse

### Properties

| | |
|---|---|
| Prio recon | Off |
| Load images to viewer | On |
| Inline movie | Off |
| Auto store images | On |
| Load images to stamp segments | Off |
| Load images to graphic segments | Off |
| Auto open inline display | Off |
| Auto close inline display | Off |
| Start measurement without further preparation | Off |
| Wait for user to start | Off |
| Start measurements | Single measurement |

### Routine

| | |
|---|---|
| Slice group | 1 |
| Slices | 60 |
| Dist. factor | 0 % |
| Position | L0.0 P0.6 H7.3 mm |
| Orientation | Transversal |
| Phase enc. dir. | A >> P |
| AutoAlign | --- |
| Phase oversampling | 0 % |
| FoV read | 210 mm |
| FoV phase | 100.0 % |
| Slice thickness | 1.25 mm |
| TR | 6400 ms |
| TE | 65.0 ms |
| Concatenations | 1 |
| Filter | None |
| Coil elements | AC |

### Contrast - Common

| | |
|---|---|
| TR | 6400 ms |
| TE | 65.0 ms |
| MTC | Off |
| Magn. preparation | None |
| Flip angle exc | 90 deg |
| Flip angle fat sat | 110 deg |
| Fat suppr. | Fat sat. |
| Fat sat. mode | Weak |

### Contrast - Dynamic

| | |
|---|---|
| Averaging mode | Long term |
| Reconstruction | Magnitude |
| Measurements | 1 |
| Delay in TR | 0 ms |
| Multiple series | Off |

### Resolution - Common

| | |
|---|---|
| FoV read | 210 mm |
| FoV phase | 100.0 % |
| Slice thickness | 1.25 mm |
| Base resolution | 168 |
| Phase resolution | 100 % |
| Phase partial Fourier | 6/8 |
| Interpolation | Off |

### Resolution - iPAT

| | |
|---|---|
| Accel. mode | GRAPPA |
| Accel. factor PE | 3 |

### Resolution - iPAT

| | |
|---|---|
| Ref. lines PE | 66 |
| Reference scan mode | GRE/separate |

### Resolution - Filter Image

| | |
|---|---|
| Distortion Corr. | Off |
| Prescan Normalize | Off |
| Dynamic Field Corr. | Off |

### Resolution - Filter Rawdata

| | |
|---|---|
| Raw filter | Off |
| Elliptical filter | Off |

### Geometry - Common

| | |
|---|---|
| Slice group | 1 |
| Slices | 60 |
| Dist. factor | 0 % |
| Position | L0.0 P0.6 H7.3 mm |
| Orientation | Transversal |
| Phase enc. dir. | A >> P |
| FoV read | 210 mm |
| FoV phase | 100.0 % |
| Slice thickness | 1.25 mm |
| TR | 6400 ms |
| Multi-slice mode | Interleaved |
| Series | Interleaved |
| Concatenations | 1 |

### Geometry - AutoAlign

| | |
|---|---|
| Slice group | 1 |
| Position | L0.0 P0.6 H7.3 mm |
| Orientation | Transversal |
| Phase enc. dir. | A >> P |
| AutoAlign | --- |
| Initial Position | L0.0 P0.6 H7.3 |
| L | 0.0 mm |
| P | 0.6 mm |
| H | 7.3 mm |
| Initial Rotation | 0.00 deg |
| Initial Orientation | Transversal |

### Geometry - Saturation

| | |
|---|---|
| Fat suppr. | Fat sat. |
| Fat sat. mode | Weak |
| Special sat. | None |

### Geometry - Navigator

### Geometry - Tim Planning Suite

| | |
|---|---|
| Set-n-Go Protocol | Off |
| Table position | H |
| Table position | 0 mm |
| Inline Composing | Off |

### System - Miscellaneous

| | |
|---|---|
| Positioning mode | FIX |
| Table position | H |
| Table position | 0 mm |
| MSMA | S - C - T |
| Sagittal | R >> L |

### System - Miscellaneous

| | |
|---|---|
| Coronal | A >> P |
| Transversal | F >> H |
| Coil Combine Mode | Sum of Squares |
| Matrix Optimization | Off |
| AutoAlign | --- |
| Coil Select Mode | Default |

### System - Adjustments

| | |
|---|---|
| B0 Shim mode | Brain |
| B1 Shim mode | TrueForm |
| Confirm freq. adjustment | Off |
| Assume Dominant Fat | Off |
| Assume Silicone | Off |
| Adjustment Tolerance | Auto |

### System - Adjust Volume

| | |
|---|---|
| ! Position | R0.9 P9.8 H15.0 mm |
| ! Orientation | T > C-16.4 > S-1.3 |
| ! Rotation | 2.64 deg |
| ! A >> P | 163 mm |
| ! R >> L | 160 mm |
| ! F >> H | 111 mm |
| Reset | Off |

### System - pTx Volumes

| | |
|---|---|
| B1 Shim mode | TrueForm |
| Excitation | Standard |

### System - Tx/Rx

| | |
|---|---|
| Frequency 1H | 297.231131 MHz |
| Correction factor | 1 |
| Gain | High |
| Img. Scale Cor. | 1.000 |
| Reset | Off |
| ? Ref. amplitude 1H | 0.000 V |

### Physio - Signal1

| | |
|---|---|
| 1st Signal/Mode | None |
| TR | 6400 ms |
| Concatenations | 1 |

### Physio - PACE

| | |
|---|---|
| Resp. control | Off |
| Concatenations | 1 |

### Diff - Neuro

| | |
|---|---|
| Diffusion mode | Free |
| Diff. directions | 30 |
| Diffusion Scheme | Monopolar |
| Diff. weightings | 2 |
| b-value 1 | 0 s/mm² |
| b-value 2 | 1000 s/mm² |
| b-value 1 | 2 |
| b-value 2 | 1 |
| Diff. weighted images | On |
| Trace weighted images | On |
| ADC maps | On |
| FA maps | On |
| Mosaic | Off |
| Tensor | On |
| Noise level | 40 |

### Diff - Body

| | |
|---|---|
| Diffusion mode | Free |
| Diff. directions | 30 |
| Diffusion Scheme | Monopolar |
| Diff. weightings | 2 |
| b-value 1 | 0 s/mm² |
| b-value 2 | 1000 s/mm² |
| b-value 1 | 2 |
| b-value 2 | 1 |
| Diff. weighted images | On |
| Trace weighted images | On |
| ADC maps | On |
| Exponential ADC Maps | Off |
| FA maps | On |
| Invert Gray Scale | Off |
| Calculated Image | Off |
| b-Value >= | 0 s/mm² |
| Noise level | 40 |

### Diff - Composing

| | |
|---|---|
| Inline Composing | Off |
| Distortion Corr. | Off |

### Sequence - Part 1

| | |
|---|---|
| Introduction | On |
| Optimization | None |
| Multi-slice mode | Interleaved |
| Free echo spacing | Off |
| Echo spacing | 0.81 ms |
| Bandwidth | 1418 Hz/Px |

### Sequence - Part 2

| | |
|---|---|
| EPI factor | 168 |
| RF pulse type | Low SAR |
| Gradient mode | Normal |
| Excitation | Standard |

### Sequence - pTX Pulses

### Sequence - Special

| | |
|---|---|
| Use ptx pulse | On |
| Exc. pul. dur | 10300 us |
| Refoc. pul. dur | 11900 us |
| Exc. pul. phase | 90 deg |
| Refoc. pul. phase | 0 deg |
| Exc. pul. TE con. | 3120 us |
| Refoc. pul. TE con. | 2560 us |
| n(exc. pulses) | 1 |
| n(refoc. pulses) | 1 |
